# Beyond Perinatal Mood: Breadth of Psychiatric Symptoms and Delay Discounting as Correlates of Mothers’ Relational Reports from Pregnancy Through Adult Children

**DOI:** 10.64898/2026.11.04.26367299

**Authors:** Rachel DeLauder, Mandy Hebert, Sydnee Rhodes, Eva Lang, Madison Lee, Annika Riddle, Adrianys Margary, Julia C. Basso

## Abstract

Maternal mental illness affects roughly one in five mothers in the United States, yet most evidence linking it to the mother–child relationship comes from the first postpartum year and relies on self-report measures of depression, anxiety, and stress. In 613 women spanning five family contexts — pregnancy and mothers whose youngest child was aged 0–2, 3–12, 13–17, or 18 and older — we examined twelve psychological constructs as correlates of mothers’ relational reports. Eleven were associated with relational reports individually; entered together, only three retained independent associations surviving false discovery rate correction: cross-domain psychiatric symptom burden (β = −0.38), delay discounting (β = −0.11), and orienting sensitivity (β = +0.10). To our knowledge, this is the first peer-reviewed study linking delay discounting to maternal relational reports. Depression, anxiety, anger, perceived stress, and negative affect fell to non-significance, and the symptom composite that displaced them contains none of those domains, indicating that breadth of psychiatric symptoms, not their type, tracked relational reports. A person-centered analysis converged: the latent profile with the poorest relational reports was defined by symptoms across multiple domains rather than by elevated stress alone. Associations varied across family contexts, though because each context used a different relational instrument, differences cannot be attributed to child age. These cross-sectional findings suggest that the psychological dimension most closely tied to maternal relational reports is broader than the perinatal mood constructs around which research and screening are organized, and that cross-domain symptom assessment warrants prospective evaluation as a complement to mood screening.

## 1.0 INTRODUCTION

Caring for a child is a sustained demand on a mother’s attention, emotion, and capacity to put the future ahead of the present. Psychiatric illness depletes all three. Maternal mental health disorders are among the most common complications of childbirth, affecting approximately 20% of women in the United States, with meta-analytic estimates for perinatal depression nearly one in seven(Gavin et al. 2005). Unfortunately, maternal mental health disorders are becoming more common: between 2016 and 2023, the proportion of US mothers rating their mental health as fair or poor rose by nearly 65%(Daw et al. 2025). In the most recent multistate review of maternal deaths, mental health conditions — including suicide and overdose — surpassed hemorrhage and cardiac conditions as the leading underlying cause of pregnancy-related death(Trost et al. 2022). Yet, detection and treatment remain poor at every step, from screening through diagnosis to remission(Cox et al. 2016).

Maternal mental illness also carries consequences across generations. Offspring of mothers with perinatal depression show increased odds of depression (OR = 1.70)(Tirumalaraju et al. 2020), disruptive behavioral disorder symptoms (OR = 1.47)(Tusa et al. 2025), and socioemotional and behavioral difficulties (OR = 1.79)(Madigan et al. 2018). These are not transient effects confined to infancy; they appear in school performance, peer relationships, emotional regulation, and the child’s own risk of psychiatric disorder into adulthood.

Maternal mental health shapes child development largely through the dyadic processes that organize early caregiving. Mothers and infants coordinate their behavior from the first weeks of life, matching gaze, vocalization, affect, and movement in exchanges that recur hundreds of times a day during feeding, soothing, and play(Feldman 2007). These exchanges are not merely social; they are the means by which an infant’s physiological and emotional states are regulated by another person before the capacity for self-regulation develops. Furthermore, they extend to the coordination of heart rate, cortisol, and autonomic arousal between mother and child(Bell 2020). Maternal depression and anxiety disrupt this coordination, reducing responsiveness to infant cues, flattening affective exchange, and desynchronizing the timing upon which co-regulation depends(A. Stein et al. 2014). The association is well documented: a meta-analysis drawing on 99 samples and 110,968 mothers found that maternal depression, anxiety, and stress were each related to poorer self-reported bonding(O’Dea et al. 2023). Importantly, the relationship between maternal mental health and responsiveness is not only a conduit for risk but also a point of intervention: programs targeting maternal sensitivity and mother–infant interaction improve attachment outcomes, with the most focused and behaviorally specific interventions proving the most effective(Bakermans-Kranenburg et al. 2003). The mother’s state and the child’s outcome are therefore linked by a relationship that is itself measurable and, in principle, modifiable — which is what makes this a dyadic problem rather than an individual one, and what makes the quality of that relationship worth measuring directly. This literature establishes that distress and bonding difficulties co-occur. What it has not settled falls into four areas: construct heterogeneity, developmental range, variable measurement, and analytic convention.

The first area is construct heterogeneity. “Attachment” is often used as an umbrella term for three distinct relational constructs: first, maternal–fetal attachment and mother–infant bonding, which concerns a mother’s feelings toward her fetus or infant; second, infant attachment security, which concerns the child’s behavior toward a caregiver; and third, a parent’s report of closeness, conflict, or communication with an older child, which is distinct yet again(Bicking Kinsey and Hupcey 2013). Reviews of parent-report measures find uneven psychometric support across instruments: of 17 self-report measures of the parent–infant bond, only one demonstrated sufficient structural validity, internal consistency and reliability from high-quality evidence, and its factor structure has not been consistently replicated(Wittkowski et al. 2020). The informant matters as well: in a cohort of 454 women followed from pregnancy to three months postpartum, prenatal anxiety disorders predicted self-reported bonding impairment but not observed interaction quality, while depressive symptoms predicted lower observed maternal sensitivity(Nath et al. 2019).

A second concern is developmental range. Most evidence comes from pregnancy and the first postpartum year, yet maternal depression does not recede on that schedule. In the largest US postpartum screening study, only 40.1% of depressive episodes began postpartum; 33.4% began during pregnancy and 26.5% before conception(Wisner et al. 2013). In a prospective cohort followed for four years, depressive symptoms were more prevalent at four years postpartum (14.5%) than at any point in the first eighteen months, and 40% of those cases arose in women who had reported no symptoms earlier(Woolhouse et al. 2015). Work on the mother–child relationship beyond infancy exists but is sparse and uses different outcome definitions: a meta-analysis of preschool attachment found maternal depressive symptoms modestly elevated for disorganized or controlling classifications, with the insecure-versus-secure estimate inconclusive(Badovinac et al. 2018); a six-year adolescent study found that maternal depressive symptoms prospectively predicted mothers’ reports of greater conflict and lower support(Hale et al. 2020); and a review of parent mental health and intergenerational attachment identified only 11 studies(Risi et al. 2021). The risk therefore extends well past the perinatal period, while the evidence base remains concentrated within it.

A third concern is what gets measured. Models of maternal relational functioning typically include depression, anxiety, and stress(O’Dea et al. 2023). They rarely include behavioral measures of decision making or dimensions of temperament outside negative affect. This matters because the affective measures overlap substantially. Across 20 years of the Dunedin cohort, psychiatric disorders were explained better by a single general psychopathology dimension than by separate internalizing, externalizing, and thought-disorder factors(Caspi et al. 2014). This non-specificity helps explain why multiple diagnoses share risk factors, biomarkers, and treatment responses(Caspi and Moffitt 2018). A model containing several correlated affective measures may therefore estimate one dimension several times rather than identify distinct contributors.

One such behavioral measure is delay discounting. Operationalized as the rate at which rewards lose value as a function of delay, it indexes the balance between the impulsive and executive systems that govern intertemporal choice(Bickel et al. 2018), and offers a behavioral alternative to the self-report affective measures that dominate this literature. It has been characterized as a trans-disease process spanning substance use disorder, major depressive disorder, and a range of maladaptive health behaviors(Bickel et al. 2019), and we have previously linked it to trauma symptomatology and dissociation(Basso et al. 2023). No peer-reviewed study to date has tested delay discounting alongside a broad set of overlapping psychological symptoms in relation to maternal reports of relationship quality.

Underlying all three is an analytic convention. Nearly all of this work is variable-centered, estimating the average association between one predictor and a relational outcome, yet psychological characteristics co-occur. Person-centered methods address this issue directly: perinatal studies have identified distinct bonding trajectories(de Waal et al. 2026) and profiles of co-occurring maternal mental-health symptoms(Kingston et al. 2018), but they have not been applied to the psychological characteristics that may precede and accompany relational difficulty.

We approached these questions from a resource account of caregiving set within a transdiagnostic view of psychopathology. Caregiving is a sustained demand on finite regulatory capacities — attention to a child’s cues, regulation of one’s own affect, and the repeated choice of delayed over immediate reward(Feldman 2007; A. Stein et al. 2014) — and under a transdiagnostic account, what depletes those capacities is general liability expressed across symptom domains rather than any single disorder(Caspi and Moffitt 2018). This framing generates three predictions. First, because the affective self-report measures on which this literature relies — depression, anxiety, anger, perceived stress, and negative affect — each index that same liability, they should prove largely redundant with one another: entered together, most should not retain unique associations with relational reports. Second, a measure of symptom breadth across non-affective domains should retain an association after adjustment for the affective measures, because it indexes depletion the affective scales do not capture. Third, constructs that index the regulatory capacities themselves rather than their depletion — delay discounting as a behavioral measure of future valuation, and temperamental effortful control and orienting sensitivity — should retain associations independent of symptom measures. The twelve constructs examined were selected to test these predictions: five affective self-report measures, one index of symptom breadth, and six dispositional or behavioral measures outside the affective self-report domain (effortful control, extraversion, orienting sensitivity, mindfulness, autism-spectrum traits, and delay discounting).

The present cross-sectional study tests these predictions in a sample of 613 women spanning five family contexts: pregnant women and mothers whose youngest living child was aged 0–2, 3–12, 13–17, or 18 years and older. Because the relational constructs are related but not equivalent, the analyses address within-context associations rather than group differences at the relational level. A sixth group of 214 women reporting on their own parents rather than on a child is analyzed separately as a contrast. The study had three aims. The first was to test these predictions by examining which of the twelve constructs show associations with relational reports independent of the others. The second was to estimate whether those associations hold across the five family contexts. The third was to test the same model from a person-centered direction: if relational difficulty tracks breadth of liability, the latent profile with the poorest relational reports should be defined by symptoms across domains rather than by elevated distress alone.

## 2.0 METHODS

### 2.1. Participants and Design

The retained sample comprised 840 women recruited through Amazon Mechanical Turk. The raw survey export contained 1,502 participant responses, of which 11 were flagged as spam, leaving 1,491 genuine responses; 246 did not reach the end of the survey. Of the 1,245 recorded as finished, 276 contained no item-level responses whatsoever and were excluded, leaving 969 genuine respondents; 71 of these failed an attention check, and 58 were earlier submissions by workers who completed the survey twice, for whom the later response was retained. The final retained sample was 840 (**Table 1**). Participants averaged 34.6 years of age (SD = 7.5, range 20–50). The sample identified predominantly as White/Caucasian (84.2%), with 7.9% Black/African American, 4.8% Asian, 2.3% Other, and 0.7% American Indian/Alaska Native. Educational attainment was high: 39.5% held a bachelor’s degree, 29.0% reported some college, and 23.8% held an advanced degree. Most participants were married (58.3%) or partnered (22.4%). The design was cross-sectional, with all measures administered in a single session. All participants completed informed consent (protocol #20-586) and procedures were approved by and in compliance with the Virginia Tech Institutional Review Board.

**Table 1.**
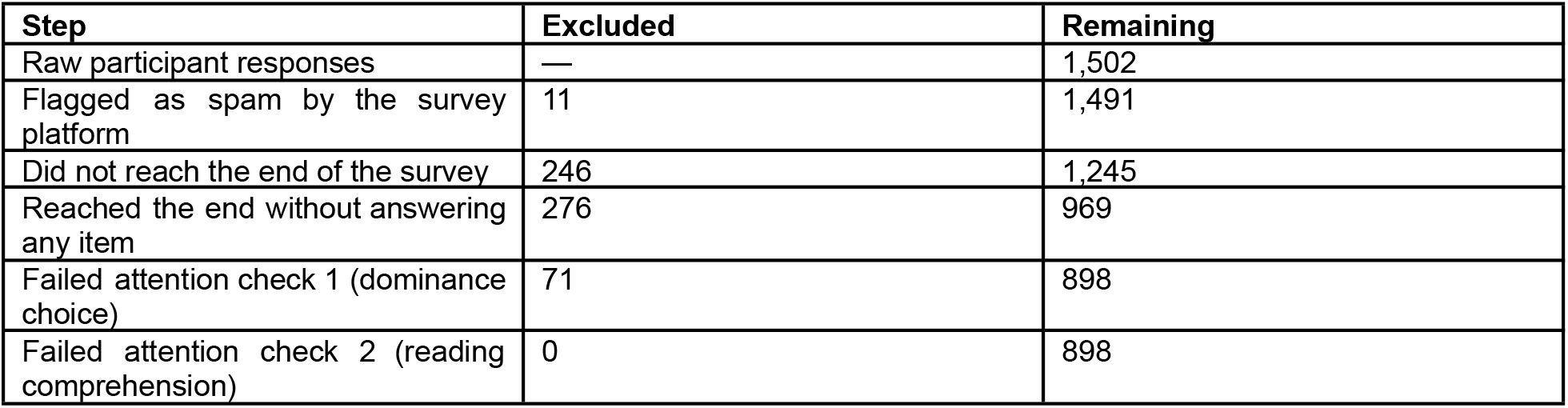
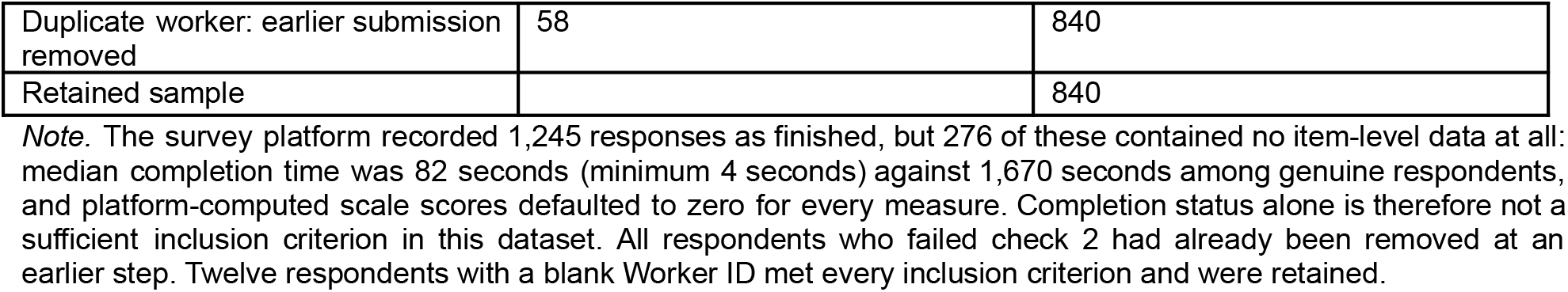
Derivation of the retained sample from the raw survey export.

| Step | Excluded | Remaining |
|---|---|---|
| Raw participant responses | — | 1,502 |
| Flagged as spam by the survey platform | 11 | 1,491 |
| Did not reach the end of the survey | 246 | 1,245 |
| Reached the end without answering any item | 276 | 969 |
| Failed attention check 1 (dominance choice) | 71 | 898 |
| Failed attention check 2 (reading comprehension) | 0 | 898 |
| Duplicate worker: earlier submission removed | 58 | 840 |
| Retained sample |  | 840 |

*Note.* The survey platform recorded 1,245 responses as finished, but 276 of these contained no item-level data at all: median completion time was 82 seconds (minimum 4 seconds) against 1,670 seconds among genuine respondents, and platform-computed scale scores defaulted to zero for every measure. Completion status alone is therefore not a sufficient inclusion criterion in this dataset. All respondents who failed check 2 had already been removed at an earlier step. Twelve respondents with a blank Worker ID met every inclusion criterion and were retained.

Participants were routed to one of six relational questionnaires based on their parenthood status and, for mothers, by the age of the youngest living child. Five of these groups reported on a fetus or child and constitute the primary analytic sample. The sixth, comprising women without children, reported on the relationship with their own parents; because this is a different relational construct with a different reporter role and relationship target, it is not pooled with the maternal-report groups and is analyzed separately in Section 3.6. All 613 women routed to a maternal instrument had a usable relational score and complete model covariates, and all contribute to the multilevel models; two lacked a PSQI global score and are excluded from the profile analyses, which use N = 611 (Table 2). Of the remaining 227, 222 were routed to the own-parents questionnaire and 5 completed no relational instrument; 8 of the 222 did not respond to either regard subscale and received no score, leaving 214. The retained sample is therefore 840 and the sample contributing to relational analyses is 827 (613 maternal-report, 214 own-parents). All parent instruments instructed participants to focus on the youngest living child, so relational scores refer to that child throughout. Routing by the youngest child aligns the relational target with the instrument validated for that age range and, for mothers of younger children, with the birth most proximal to any perinatal mood symptoms. Most mothers in the three younger groups (61.1% in the 0–2 group, 68.7% in the 3–12 group, 60.4% in the 13–17 group), and 42.5% of mothers of adult children, had more than one child, so the groups index the age of the youngest child rather than parenting experience or family size. Because each participant completed exactly one relational instrument, determined by her family context, the instrument is perfectly confounded with parenthood status and child age. We refer to this five-level variable as instrument group throughout, and adjust for it in all analyses; slope differences across groups cannot be attributed to child age alone.

**Table 2.** Characteristics of the five maternal-report groups.

| Group | Instrument | n | Age M (SD) | White % | Hispanic % | BA+ % | Partnered % |
|---|---|---|---|---|---|---|---|
| Pregnant | PAI | 39 | 27.8 (4.7) | 84.6 | 12.8 | 84.6 | 97.4 |
| Child 0–2 years | MAI | 145 | 30.7 (5.6) | 80.7 | 8.3 | 69.0 | 94.5 |
| Child 3–12 years | CPRS | 298 | 36.2 (6.5) | 86.2 | 6.4 | 58.7 | 86.9 |
| Child 13–17 years | RIPA | 91 | 42.0 (5.0) | 90.1 | 4.4 | 53.8 | 73.6 |
| Child 18+ years | RIPA-AC | 40 | 45.4 (3.7) | 90.0 | 0.0 | 52.5 | 72.5 |
| Total |  | 613 | 35.8 (7.5) | 85.6 | 6.5 | 61.7 | 86.5 |

*Note.* PAI = Prenatal Attachment Inventory; MAI = Maternal Attachment Inventory; CPRS = Child–Parent Relationship Scale; RIPA = Revised Inventory of Parental Attachment; RIPA-AC = RIPA, Adult Child version. BA+ = bachelor’s degree or higher. All 613 participants routed to a maternal instrument had a usable relational score. Two were missing the PSQI global score, so the profile analyses use N = 611; the multilevel models use all 613.

### 2.2. Measures

Participants completed all self-report scales while refraining from alcohol or illicit substances. Internal consistency is reported both as published and as observed in the present sample. Fifty-eight participants completed the survey twice, approximately 60 days apart, providing an incidental test–retest subsample; stability was acceptable to good for most measures (median r = .78; **Supplementary Table S1**). Full measure descriptions, including the six relational instruments, are given in Supplementary Methods.

#### 2.2.1. Relational measures

Five maternal-report instruments were administered, one per group: the Prenatal Attachment Inventory (PAI; 21 items; α = .91)(Muller 1993), the Maternal Attachment Inventory (MAI; 26 items; α = .98)(Müller 1994), the Child–Parent Relationship Scale (CPRS; 30 items across conflict, positive aspects, and dependence subscales; α = .80)(Driscoll and Pianta 2011), the Revised Inventory of Parental Attachment (RIPA; 30 items; α = .75)(Johnson et al. 2003), and an adult-child version of the RIPA (α = .44). The published RIPA validation (N = 212 parents) reported α = .91 for trust/avoidance and α = .72 for communication; the adult-child version is an adaptation for which no separate published validation exists, and findings for that group warrant caution. The MAI showed a pronounced ceiling effect: 34 of 145 mothers (23.4%) selected the identical response to every item, 33 of them the maximum. All MAI items are positively worded and the instrument contains no conflict or difficulty items, which plausibly accounts for both the ceiling effect and the inflated reliability estimate. Straight-lining was absent (0%) for the CPRS, RIPA, and PAI. Women without children completed the Parent–Adult Child Relationship Questionnaire (PACRQ; 13 mother items, α = .86; 13 father items, α = .70)(Peisah et al. 2019), reporting on the relationship with their own parents. The relational score for this group was the mean of the standardized Mother and Father Regard subscales, requiring at least one; the Responsibility and Control subscales index filial obligation and parental dominance rather than attachment and were not included.

#### 2.2.2. Psychological measures

The DSM-5 Level 1 Cross-Cutting Symptom Measure (23 items; α = .95) yields 13 domain scores using the standard highest-item-within-domain rule(American Psychiatric Association n.d.). The measure is designed for domain-level rather than total scoring; validation studies of translated adult versions report total-score alphas of .94 and .95. The cross-domain psychiatric symptom burden composite used throughout is the mean of eight standardized domain scores — somatic symptoms, suicidal ideation, psychosis, memory, repetitive thoughts and behaviors, dissociation, mania, and substance use (α = .91). Depression, anxiety, and anger were excluded from the composite and entered as separate predictors so that affective symptoms could be adjusted for directly; sleep was excluded because it was measured separately by the PSQI, and personality functioning because it is not a symptom domain. A distress composite was formed as the mean of four standardized scores — perceived stress, negative affect, and reverse-scored mindfulness and effortful control (α = .79) — on the basis of the factor structure identified in preliminary analyses. It is used only in Model B. The Mindful Attention Awareness Scale (MAAS; 15 items; α = .93)(Brown and Ryan 2003), the Perceived Stress Scale (PSS; 10 items; α = .89)(Cohen et al. 1983), the Adult Temperament Questionnaire (ATQ) short form (77 items; α = .91; four higher-order factors and thirteen lower-order scales)(Evans and Rothbart 2007), and the Autism Spectrum Quotient (ASQ; 50 items; α = .86, used dimensionally)(Baron-Cohen et al. 2001) were also administered. Delay discounting was assessed with the 5-Trial Adjusting Delay Discounting Task, also known as the Minute Discounting Task(Koffarnus and Bickel 2014). Participants made five successive choices between $500 now and $1,000 after a delay, with the delay adjusted after each choice. The final delay yields the delay at which the delayed reward is valued at half its nominal amount (ED50), from which the discount rate k is derived; higher ln(k) indicates steeper devaluation of delayed rewards, that is, greater choice impulsivity. The 60-day test–retest correlation in the present sample was r = .64.

#### 2.2.3. Health measures

The Pittsburgh Sleep Quality Index(Azmeh Shahid et al. 2011), a 41-item medical history checklist(PhenX Toolkit 2006), the Stanford Brief Activity Survey(Taylor-Piliae et al. 2005), a sitting/sedentary behavior protocol(Marshall et al. 2010), and the Block rapid food screeners for dietary fat and fruit and vegetable intake(Block et al. 2000) were administered. Body mass index was computed from self-reported height and weight. PSQI scoring in this dataset required correction; see Supplementary Methods.

### 2.3. Derivation of the Relational Score

The five instruments are on incommensurable raw metrics, with group means ranging from 43.1 to 89.8 and standard deviations from 5.4 to 19.3, so a one-unit change does not carry the same meaning across groups. Scores were therefore standardized within the subsample completing each instrument. For the four instruments yielding a total score (PAI, MAI, RIPA, RIPA-AC), the standardized total was used. The CPRS has no total score, so its three subscales were signed so that higher values indicate a more positive relational report (Conflict and Dependence reverse-scored), their z-scores averaged, and the result re-standardized. Results are unchanged under a simpler signed-total version of the CPRS score, which correlates .97 with the composite; across the whole sample the two versions of the outcome correlate .98 and yield symptom burden slopes of −0.39 and −0.40.

#### 2.3.1. Interpretive constraint

Because standardization is within an instrument, every group has a mean of zero by construction. The score supports analysis of correlates within and across groups but cannot test group differences at the relational level, and no such comparison is reported.

### 2.4. Analytic Strategy

Analyses proceeded in four stages. Because the five groups differ greatly in size (39 to 298), we used a Bayesian multilevel model with partial pooling as the primary analysis, allowing all groups to be retained.

First, all twelve psychological constructs were examined as correlates of relational scores, entered singly (adjusting for group) and then simultaneously, with variance inflation factors reported. Influence diagnostics (Cook’s distance), Huber robust regression, and a 2,000-resample percentile bootstrap were applied to the pooled joint model as sensitivity analyses, together with re-estimation excluding respondents who answered every item of the Maternal Attachment Inventory identically. Second, a Bayesian multilevel model was fitted with instrument group as a random factor and group-varying slopes for the constructs retaining independent associations, together with demographic covariates (age, education, household income, household size, partnership status, smoking). Priors were weakly informative: Normal(0, 0.5) for regression effects, HalfNormal(0, 0.3) for group-level standard deviations, HalfNormal(0, 1) for residual error, with a non-centered parameterization. Missing responses on smoking (n = 3) and partnership status were coded as absent. Four chains of 2,000 post-warmup draws were run after 2,000 tuning iterations with target acceptance of .95. An initial Model A fit produced one divergent transition; the model was refitted with 3,000 draws after 3,000 tuning iterations at target acceptance of .99, which produced no divergences and estimates agreeing to three decimal places, and the reported Model A estimates are from that fit. Third, latent profile analysis estimated profiles from seven standardized indicators entered without reverse-coding — perceived stress, mindfulness, negative affect, effortful control, cross-domain psychiatric symptom burden, sleep problems, and medical burden — using Gaussian mixture models with diagonal covariance and 30 random starts, with solutions from one to six classes compared on BIC, entropy, smallest-class size, and bootstrapped likelihood ratio tests. Higher values therefore indicate worse functioning for five indicators and better functioning for mindfulness and effortful control. Profile differences were tested by modal assignment, by the Bolck–Croon–Hagenaars correction for classification error, and in a 30% hold-out subsample. Fourth, robustness was assessed with a specification curve estimating the focal symptom–relationship association across 144 defensible specifications of a single estimand within the maternal-report groups, crossing symptom definition, sleep covariate, covariate set, composite versus individual affective controls, group fixed effect, and exclusion of MAI straight-lining respondents.

Analyses were conducted in Python 3.12 using statsmodels, scikit-learn, PyMC, and ArviZ.

#### 2.4.1. Analytic decisions made after data inspection

The predictions stated in the Introduction were formulated before analysis but were not preregistered. Five decisions were made after inspecting the data and are reported as post hoc rather than prespecified. First, twelve respondents with blank Worker IDs were retained after it was established that they met all other inclusion criteria. Second, the primary analysis was restricted to the five maternal-report groups because the sixth group reported on a different relational construct with a different reporter role and relationship target. Third, health behaviors were dropped as a predictor block after they failed to add incrementally to the psychological constructs; they were retained as indicators in the profile analysis, which was specified separately. Fourth, the examination of lower-order temperament scales reported in Section 3.5 was prompted by the pattern of results and is labeled exploratory throughout. Fifth, the distress composite was derived from preliminary factor analyses of this dataset conducted before relational outcomes were examined, but Model B, which gives it group-varying slopes, was not specified a priori and is reported as a follow-up. The specification curve in Section 3.4 quantifies analytic flexibility for the focal association but not for the exploratory analyses.

## 3.0 RESULTS

### 3.1. Psychological Correlates of Relational Scores

Twelve psychological constructs were examined. Entered singly and adjusting only for instrument group, eleven of the twelve showed associations surviving false discovery rate (FDR) correction (**Table 3**). Entered together, only three remained statistically associated after adjustment for the other measured constructs: cross-domain psychiatric symptom burden, delay discounting, and orienting sensitivity, all three surviving false discovery rate correction across the twelve tests (p-FDR < .001, .033, and .039; sensitivity analyses in Section 3.4). Depression, anxiety, anger, perceived stress, negative affect, effortful control, mindfulness, and autism-spectrum traits were each substantially associated on their own and each reduced to non-significance in the joint model. Variance inflation factors indicate why: the DSM-5 affective domains and perceived stress carry variance inflation factors of 2.55 to 3.72, while delay discounting and orienting sensitivity carry 1.27 and 1.24, the lowest in the set. Because the composite reflects both the number of affected domains and severity within them, a sensitivity analysis replaced it with a count of domains endorsed at or above the DSM-5 Level 1 threshold (r = .97 with the composite). The count carried a joint coefficient of β = −0.286, 95% CI [−0.405, −0.166], p < .001, supporting an interpretation in terms of breadth of symptom endorsement, although the severity-weighted composite explained somewhat more variance (R² = .225 against .208). Replacing the composite and the three separately entered affective domains with the full 13-domain total reproduced the pooled association (β = −0.288, p < .001) but explained less variance (R² = .207 against .225) and cannot distinguish breadth from affective symptom load; entering the total alongside the affective domains inflates collinearity (VIF = 5.71) and was not pursued. A three-factor confirmatory model fitted to establish that the predictor set has coherent latent structure is reported in **Supplementary Table S7**.

**Table 3.**
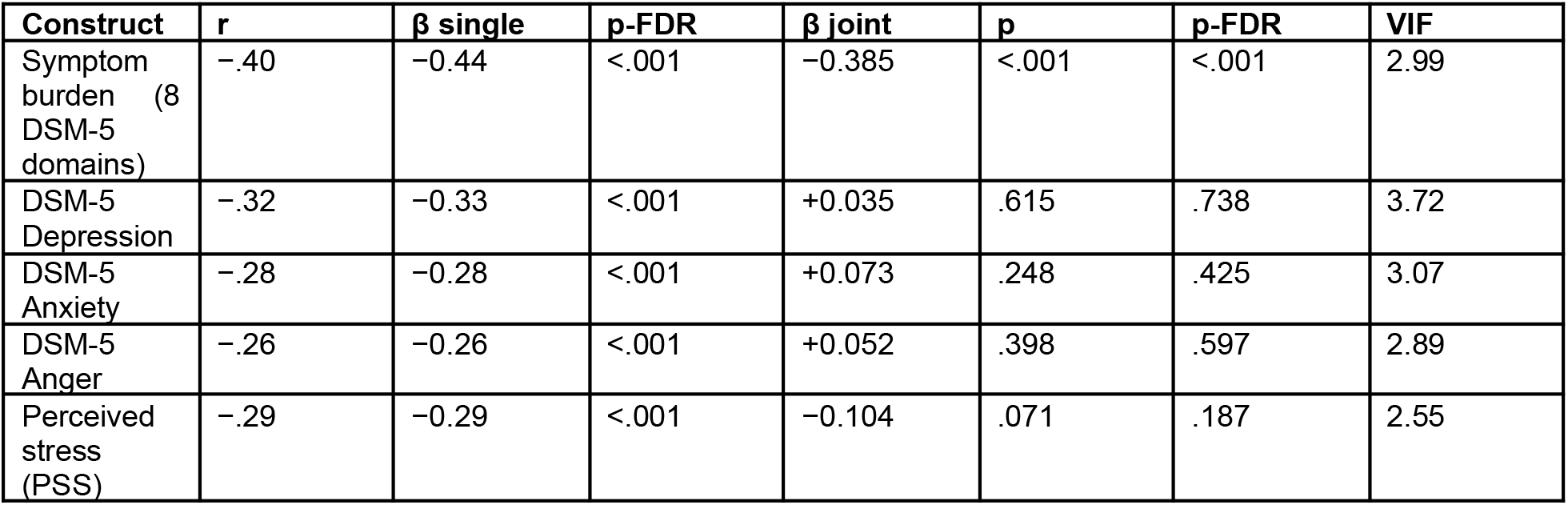
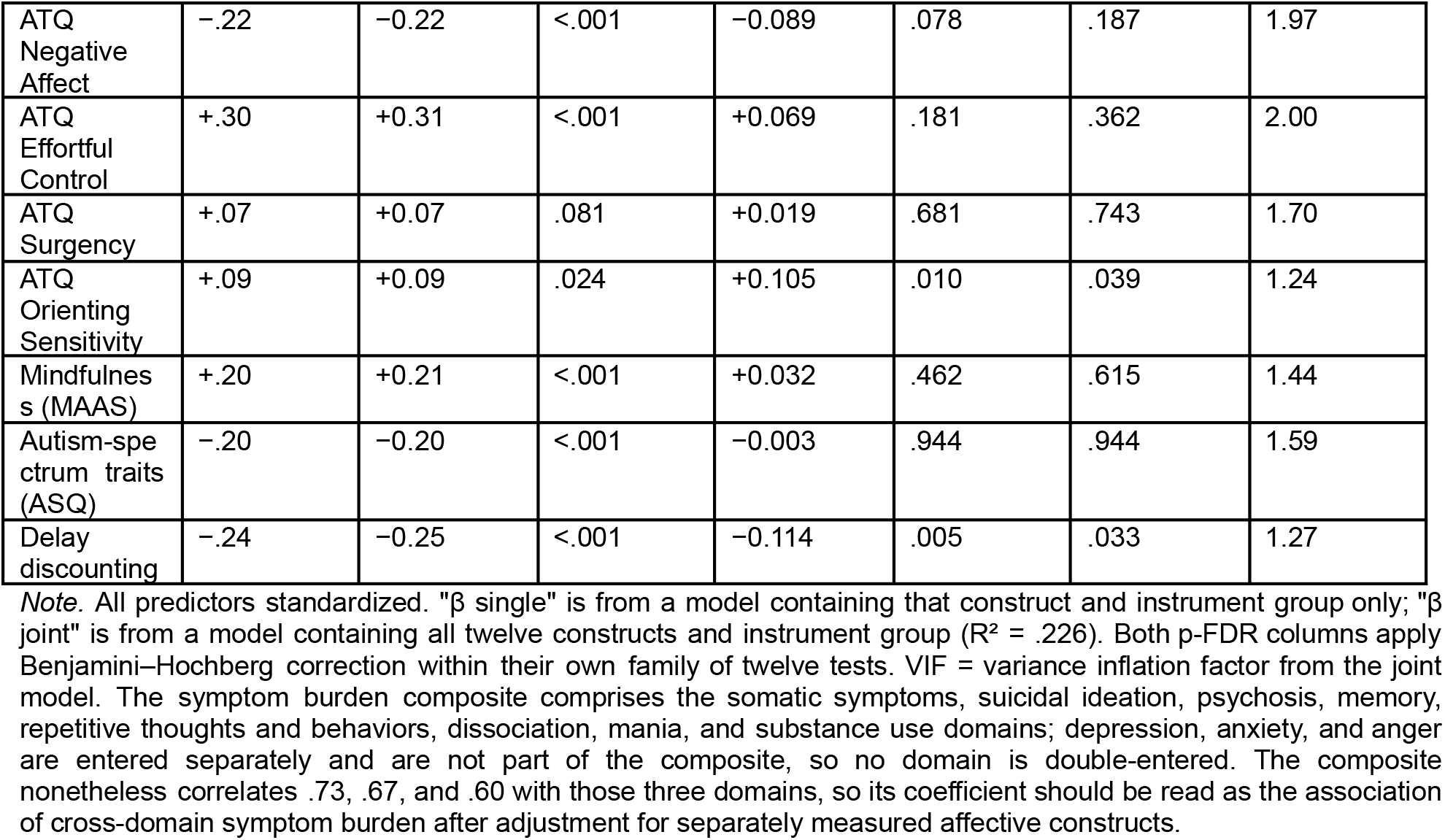
Twelve psychological constructs as correlates of relational scores, entered singly and simultaneously (N = 613).

| **Construct** | **r** | **β single** | **p-FDR** | **β joint** | **p** | **p-FDR** | **VIF** |
|---|---|---|---|---|---|---|---|
| Symptom burden (8 DSM-5 domains) | −.40 | −0.44 | <.001 | −0.385 | <.001 | <.001 | 2.99 |
| DSM-5 Depression | −.32 | −0.33 | <.001 | +0.035 | .615 | .738 | 3.72 |
| DSM-5 Anxiety | −.28 | −0.28 | <.001 | +0.073 | .248 | .425 | 3.07 |
| DSM-5 Anger | −.26 | −0.26 | <.001 | +0.052 | .398 | .597 | 2.89 |
| Perceived stress (PSS) | −.29 | −0.29 | <.001 | −0.104 | .071 | .187 | 2.55 |
| ATQ Negative Affect | −.22 | −0.22 | <.001 | −0.089 | .078 | .187 | 1.97 |
| ATQ Effortful Control | +.30 | +0.31 | <.001 | +0.069 | .181 | .362 | 2.00 |
| ATQ Surgency | +.07 | +0.07 | .081 | +0.019 | .681 | .743 | 1.70 |
| ATQ Orienting Sensitivity | +.09 | +0.09 | .024 | +0.105 | .010 | .039 | 1.24 |
| Mindfulness (MAAS) | +.20 | +0.21 | <.001 | +0.032 | .462 | .615 | 1.44 |
| Autism-spectrum traits (ASQ) | −.20 | −0.20 | <.001 | −0.003 | .944 | .944 | 1.59 |
| Delay discounting | −.24 | −0.25 | <.001 | −0.114 | .005 | .033 | 1.27 |

*Note.* All predictors standardized. "β single" is from a model containing that construct and instrument group only; "β joint" is from a model containing all twelve constructs and instrument group (R² = .226). Both p-FDR columns apply Benjamini–Hochberg correction within their own family of twelve tests. VIF = variance inflation factor from the joint model. The symptom burden composite comprises the somatic symptoms, suicidal ideation, psychosis, memory, repetitive thoughts and behaviors, dissociation, mania, and substance use domains; depression, anxiety, and anger are entered separately and are not part of the composite, so no domain is double-entered. The composite nonetheless correlates .73, .67, and .60 with those three domains, so its coefficient should be read as the association of cross-domain symptom burden after adjustment for separately measured affective constructs.

### 3.2. Bayesian Multilevel Models

Two multilevel models were fitted on identical cases (N = 613). Model A carried group-varying slopes for the three constructs retaining independent associations in the joint regression: symptom burden, delay discounting, and orienting sensitivity. Model B replaced orienting sensitivity with the distress composite in order to test a reversal observed in an earlier analysis of these data, and is reported as a follow-up rather than as the primary model. Both included demographic covariates at the population level. Chains converged cleanly in both (maximum R-hat = 1.002, minimum bulk effective sample size 3,252, zero divergent transitions; Model A was fitted with target acceptance of .99 after an initial fit produced one divergence). Group-specific slopes for both models are shown in **Figure 1** and **Table 4**.

**Figure 1.**
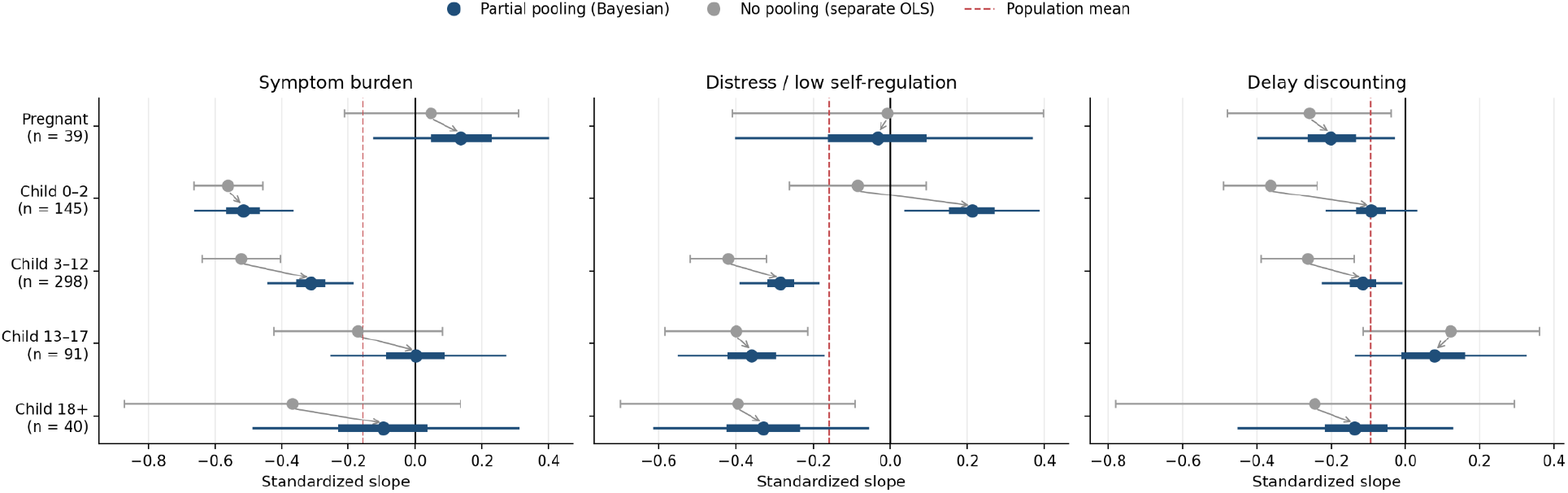
Group-specific slopes under partial pooling (N = 613). Grey markers are unpooled ordinary least squares estimates fitted separately within each group, with 95% confidence intervals; blue markers are posterior means from the Bayesian multilevel model, with thick and thin bars showing 50% and 95% credible intervals. Arrows indicate shrinkage toward the population mean (red dashed line). Panels show the three predictors of Model B, the follow-up model used to test the reversal described in Section 3.2.2; Model A estimates are given in Table 4 and Supplementary Table S2.

**Table 4.**
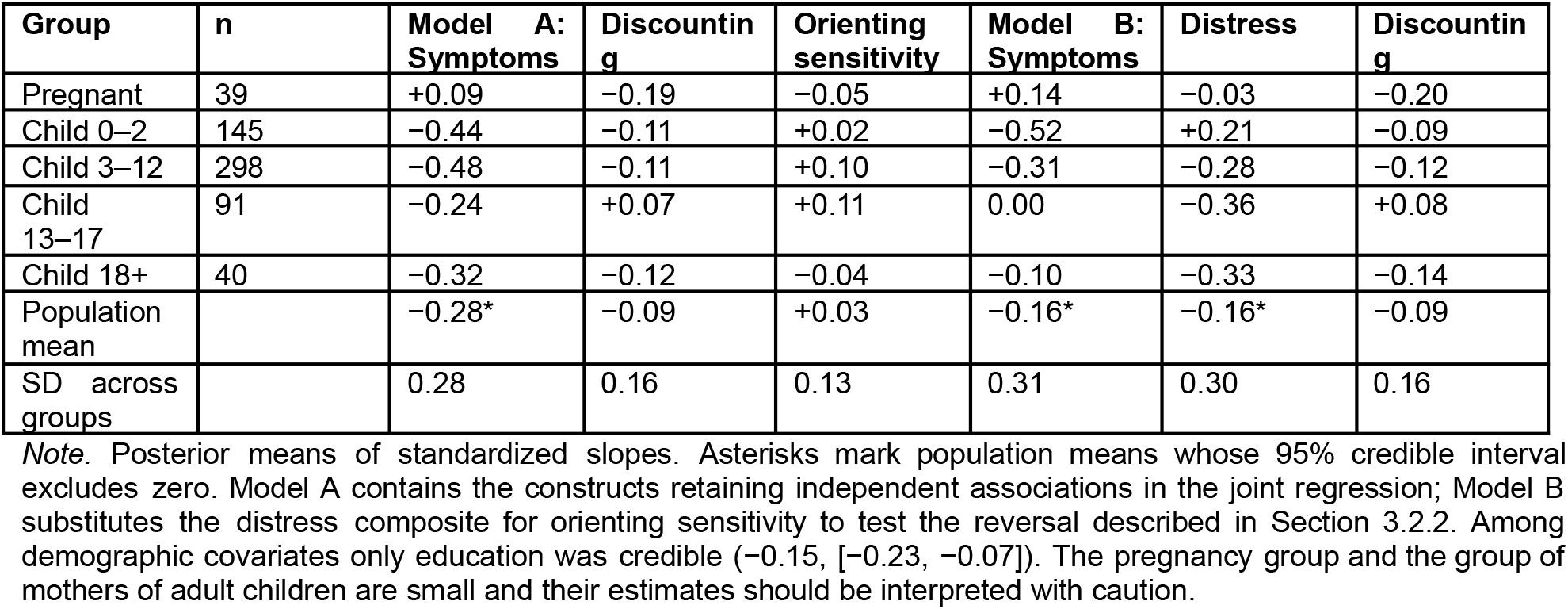
Group-specific standardized slopes from the Bayesian multilevel models (N = 613).

| Group | n | Model A: Symptoms | Discounting | Orienting sensitivity | Model B: Symptoms | Distress | Discounting |
|---|---|---|---|---|---|---|---|
| Pregnant | 39 | +0.09 | −0.19 | −0.05 | +0.14 | −0.03 | −0.20 |
| Child 0–2 | 145 | −0.44 | −0.11 | +0.02 | −0.52 | +0.21 | −0.09 |
| Child 3–12 | 298 | −0.48 | −0.11 | +0.10 | −0.31 | −0.28 | −0.12 |
| Child 13–17 | 91 | −0.24 | +0.07 | +0.11 | 0.00 | −0.36 | +0.08 |
| Child 18+ | 40 | −0.32 | −0.12 | −0.04 | −0.10 | −0.33 | −0.14 |
| Population mean |  | −0.28* | −0.09 | +0.03 | −0.16* | −0.16* | −0.09 |
| SD across groups |  | 0.28 | 0.16 | 0.13 | 0.31 | 0.30 | 0.16 |

*Note.* Posterior means of standardized slopes. Asterisks mark population means whose 95% credible interval excludes zero. Model A contains the constructs retaining independent associations in the joint regression; Model B substitutes the distress composite for orienting sensitivity to test the reversal described in Section 3.2.2. Among demographic covariates only education was credible (−0.15, [−0.23, −0.07]). The pregnancy group and the group of mothers of adult children are small and their estimates should be interpreted with caution.

#### 3.2.1. Model A

Symptom burden was credibly negative at the population level (−0.28, 95% CrI [−0.39, −0.17]) and credibly negative in the two youngest child-age groups (child 3–12, −0.48 [−0.60, −0.36]; child 0–2, −0.44 [−0.58, −0.31]). Delay discounting was negative at the population level with an interval marginally including zero (−0.09, [−0.19, 0.01]) and credibly negative among pregnant women (−0.19 [−0.39, −0.02]) and mothers of children aged 3–12 (−0.11 [−0.22, −0.00]). Orienting sensitivity was not credible at the population level (+0.03, [−0.09, 0.13]) and credible only among mothers of children aged 3–12 (+0.10 [+0.01, +0.19]), indicating that its contribution in the pooled regression is confined to one group once slopes are allowed to vary. Group-specific credible intervals are given in **Supplementary Table S2**.

#### 3.2.2. Model B and the reversal among mothers of infants

With the distress composite entered in place of orienting sensitivity, the distress slope was positive among mothers of children aged 0–2 (+0.21, 95% CrI [+0.04, +0.38]; posterior probability .992 of being positive) and negative in all three older child-age groups (−0.28, −0.36, and −0.33). The contrast between the infant-group and child 3–12 slopes was +0.50, 95% CrI [+0.30, +0.69], with posterior probability > .999 of exceeding zero, so the two slopes differ credibly rather than merely falling on opposite sides of zero. The infant-group slope also exceeded the mean of the three older child-age groups (+0.54, 95% CrI [+0.32, +0.75]) but did not credibly differ from the pregnancy group (+0.25, 95% CrI [−0.17, +0.66]), whose slope was near zero and imprecisely estimated. Partial pooling shrinks each group’s estimate toward the population mean in proportion to its imprecision and therefore works against extreme group estimates, yet the reversal persists under shrinkage. It does not, however, address the confounding of instrument with group: the reversal may reflect the relational context, the content of the Maternal Attachment Inventory, or both. That instrument contains no conflict or difficulty items and showed straight-lining in 23.4% of respondents, while the other instruments showed none; the reversal was unchanged, and if anything strengthened, when those respondents were excluded (Section 3.4.2). Entering distress alongside symptom burden also redistributes variance between two correlated constructs, so the symptom slopes in Model B differ from those in Model A and should not be read as independent estimates.

#### 3.2.3. Common-slope and varying-slope estimates answer different questions

The joint regression (**Table 3**) and the specification curve estimate a single common slope pooled across groups; the multilevel models estimate five group-specific slopes and summarize their mean. These are not the same quantity, and the paper reports both. Fitted on identical cases, predictors, and covariates, the common-slope estimate for symptom burden was −0.38 (95% CI [−0.47, −0.30]) in a Model A specification and −0.28 ([−0.37, −0.18]) in a Model B specification, against varying-slope population means of −0.28 and −0.16 respectively. For delay discounting the corresponding values were −0.10 ([−0.18, −0.03]) and −0.12 ([−0.19, −0.04]) against population means of −0.09 and −0.09. The common-slope estimates are larger and more precise because they borrow strength across groups while assuming a shared effect; the varying-slope population means are attenuated and more uncertain because group-level variation is substantial (SD = 0.28–0.31 for symptom burden). The specification curve should accordingly be read as robustness evidence for the pooled common-slope association, not as evidence that the association is negative within every family context.

### 3.3. Latent Profile Analysis

A three-profile solution was retained (N = 611, entropy = .860, mean posterior probability = .937); class enumeration indices for one- to six-class solutions are given in **Supplementary Table S3**. Profile 1 (n = 96) was defined by psychiatric symptom burden 1.77 SD above the mean with elevated medical burden (1.32), poor sleep, and low effortful control. Profile 2 (n = 282) showed elevated stress and negative emotionality with symptom burden near the sample mean. Profile 3 (n = 233) showed the inverse pattern across all seven indicators (Figure 2; indicator means in **Supplementary Table S4**). The labels distressed–symptomatic, mildly strained, and resilient describe relative standing on the indicators and do not imply clinical status or demonstrated resilience.

**Figure 2.**
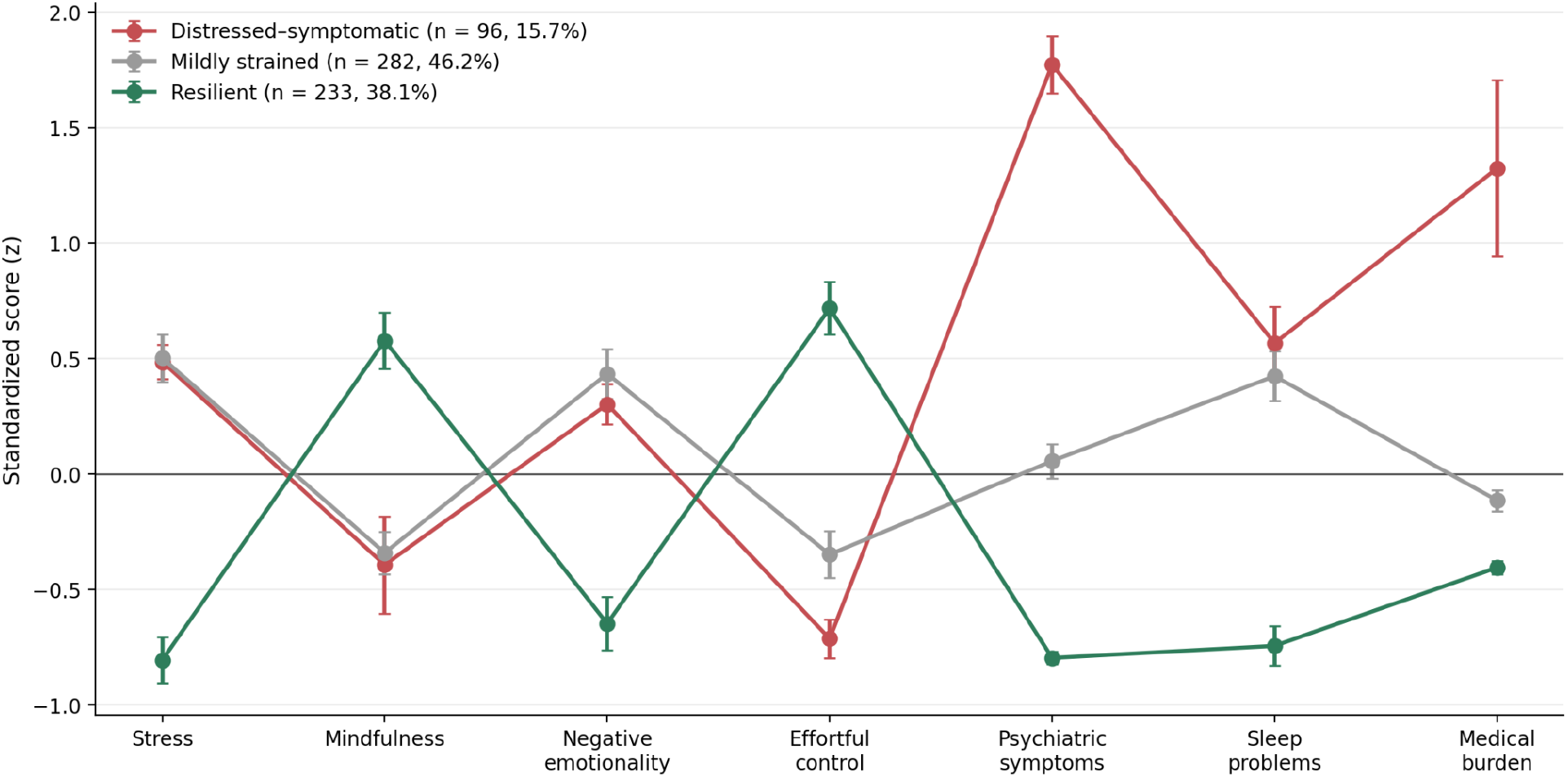
Latent profiles of psychological and health functioning (N = 611). Points are profile means on each standardized indicator; error bars are 95% confidence intervals. The distressed–symptomatic profile is distinguished primarily by psychiatric symptom burden and medical burden; the resilient profile shows the inverse pattern across all seven indicators.

Relational scores differed across profiles in the expected ordering, F(2, 608) = 63.73, p < .001, η² = .173, with means of −0.86, −0.02, and +0.38 (**Figure 3**). All three pairwise contrasts survived false discovery rate correction (d = 1.31, 0.84, and 0.46). Profile prevalence differed across instrument groups, ranging from 51.3% of pregnant women to 0% of mothers of adult children in the distressed–symptomatic profile, so the association was re-estimated adjusting for group: F(2, 604) = 70.23, p < .001, partial η² = .189. The profile differences are therefore not attributable to the uneven distribution of profiles across family contexts.

**Figure 3.**
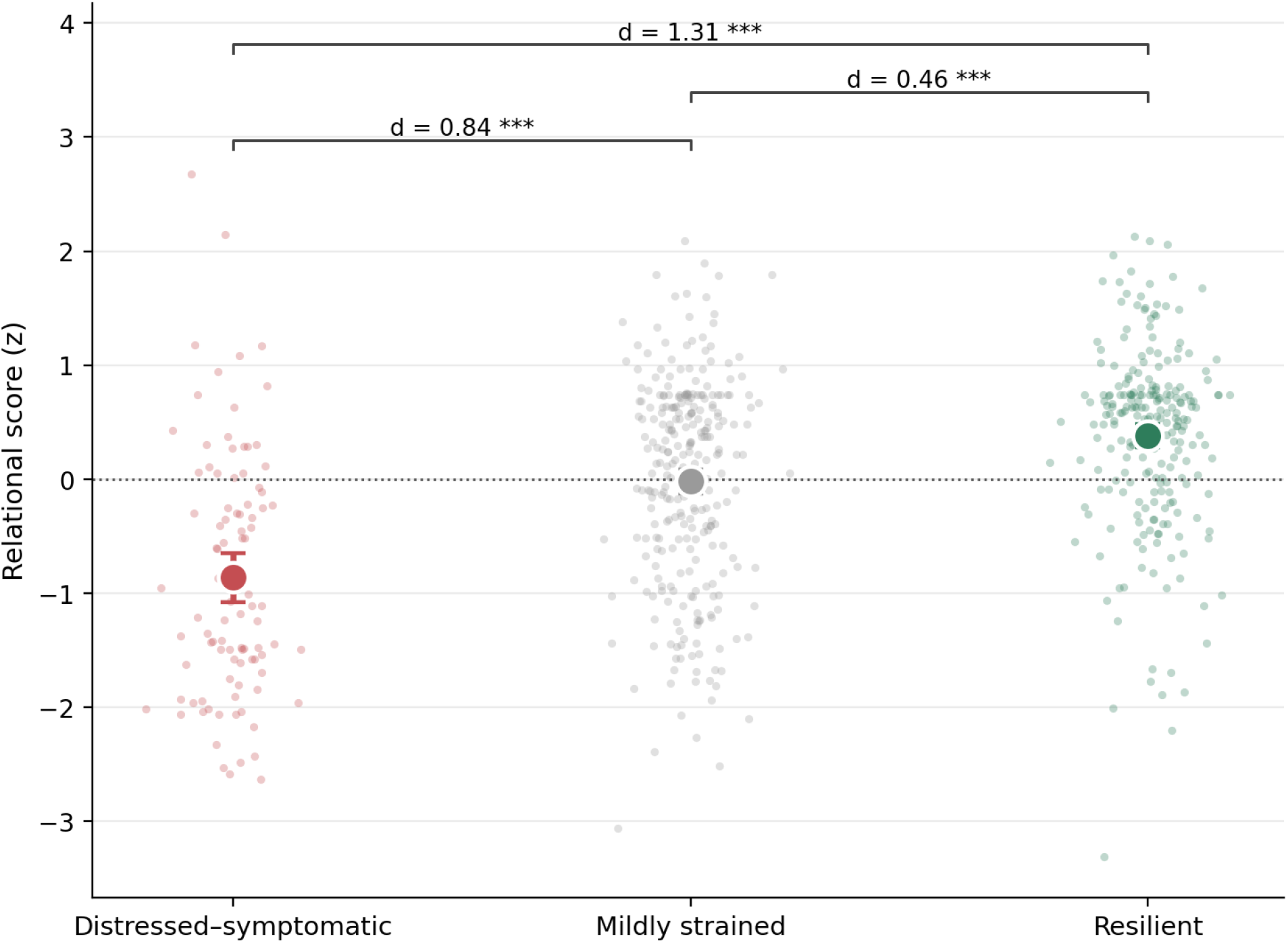
Relational scores by latent profile (N = 611). Small points are individual participants; large markers are profile means with 95% confidence intervals. Brackets give Cohen’s d for each pairwise contrast; all three survived false discovery rate correction. *** p < .001.

#### 3.3.1. Correction for classification error

Classification was accurate, with diagonal entries of the error matrix between .92 and .95. Applying the Bolck–Croon–Hagenaars correction gave class means of −0.88, −0.04, and +0.40 (SE 0.11, 0.06, 0.05), Wald χ²(2) = 150.31, p < .001 (**Supplementary Table S5**). The distal outcome findings are therefore not attributable to misclassification.

#### 3.3.2. Hold-out validation

The sample was split 70/30, stratified by instrument group, with profiles estimated in the training set (n = 427) and held-out cases (n = 184) assigned by the training model. The profile centroids were highly similar between the training and held-out samples (r = .989). Relational scores differed across profiles in the held-out set, F(2, 181) = 7.86, p < .001, η² = .080, with means of −0.34, −0.03, and +0.47 preserving the ordering. The effect is attenuated relative to the full-sample estimate (η² = .173 to .080). We report this as an attenuated internal replication rather than attributing the attenuation to test-set size: the ordering is preserved and the association remains clearly non-null, but its magnitude out of sample is uncertain. Because the profile solution was estimated in the same dataset examined throughout, this is an internal robustness check rather than independent confirmation.

#### 3.3.3. Sensitivity to the medical burden indicator

Medical burden is one of the seven profile indicators and the second strongest feature of the distressed–symptomatic profile (1.32 SD above the mean). It also showed the weakest test–retest reliability of the measures assessed (**Supplementary Table S1**), reflecting a small number of respondents who endorsed nearly all conditions at one administration and none at the other. Re-estimating the solution without medical burden produced a poorer-fitting model: entropy fell from .860 to .791 and the association with relational scores from η² = .173 to .128. The distressed–symptomatic profile was unaffected, retaining 99% of its members, while the boundary between the two remaining profiles shifted. The three-profile solution was itself fully stable across random starts. Medical burden therefore contributes signal to the separation of the two lower-symptom profiles despite its unreliability, and the solution would likely be sharper with a validated comorbidity index.

### 3.4. Robustness

The symptom–relationship association was estimated across 144 defensible specifications within the maternal-report groups, crossing symptom definition, sleep covariate, covariate set, composite versus individual affective controls, group fixed effect, and exclusion of the 34 MAI straight-lining respondents. The effect was negative in all 144 specifications and reached p < .05 in all 144 (**Figure 4)**, with a median standardized coefficient of −0.290, interquartile range [−0.317, −0.271], and full range [−0.379, −0.213]. Excluding straight-lining respondents shifted the median from −0.310 to −0.280, the second-smallest effect of any specification dimension. Group inclusion was deliberately not varied, because adding the own-parent group changes the relational construct under study rather than the analytic specification of a single estimand. The conventional specification-curve display, showing which analytic choices produced each estimate, is given in **Supplementary Figure S1**.

**Figure 4.**
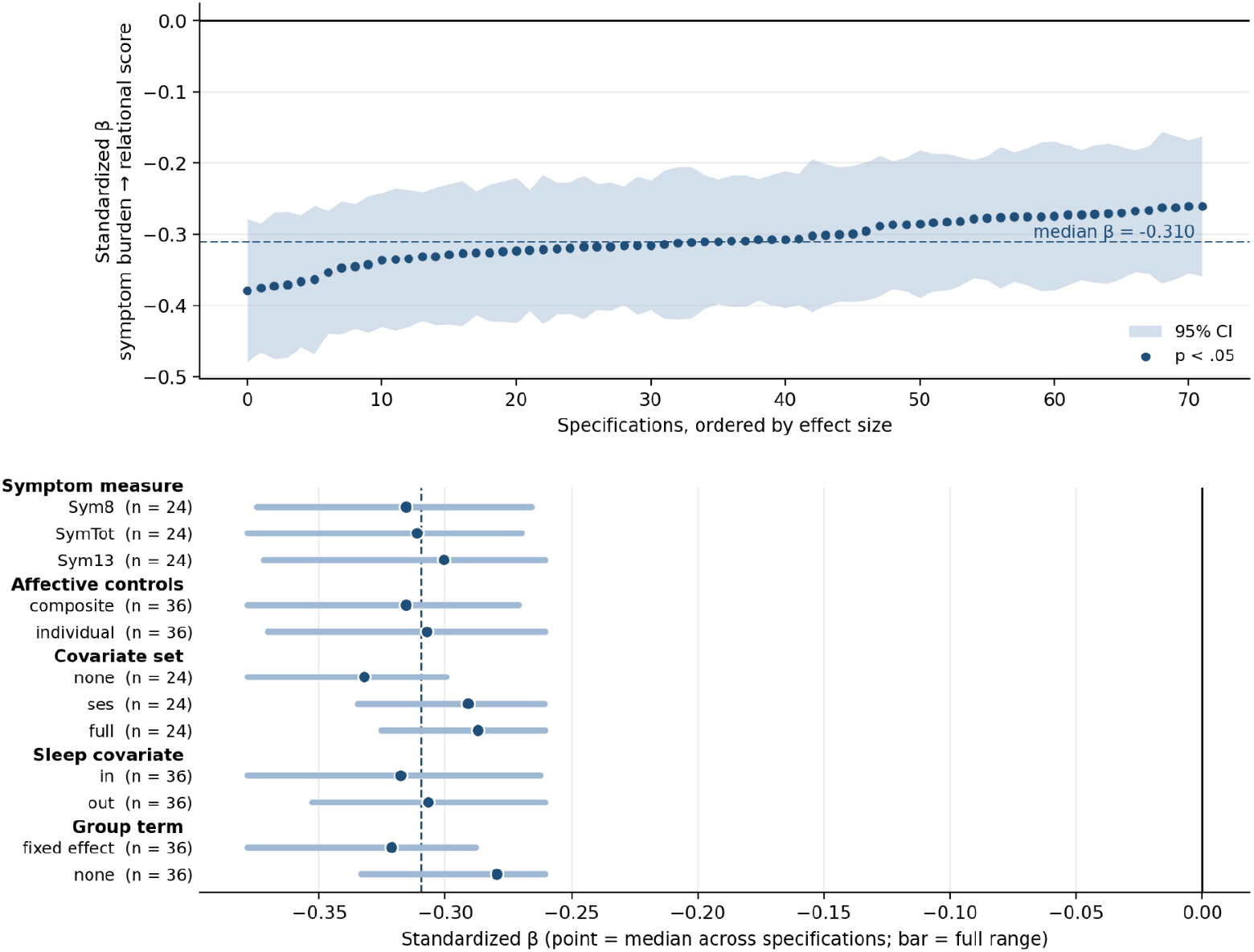
Specification curve for the association between symptom burden and relational scores. Upper panel: standardized estimates from 144 analytic specifications within the maternal-report groups, ordered by effect size, with 95% confidence intervals; all 144 were negative and all reached p < .05. Lower panel: median estimate (point) and full range (bar) for each analytic option. The dashed line marks the overall median.

#### 3.4.1. Health behaviors showed no clear incremental contribution

Dietary fat intake, fruit and vegetable intake, sleep quality, physical activity, sedentary time, body mass index, and medical burden were entered as a block after symptom burden, distress, and delay discounting. The block did not improve fit, F(7, 596) = 1.76, p = .094, and ΔR² = .016 (**Supplementary Table S6**). Zero-order associations present for dietary fat (r = −.23), medical burden (−.28), sleep (−.19), and fruit and vegetable intake (−.15) were reduced to non-significance on adjustment, and the sleep coefficient reversed sign. Physical activity showed no association at any stage (r = −.01). There was therefore no clear incremental contribution from these health behaviors after adjustment for symptom burden, distress, and delay discounting, and they are not retained. These data do not identify which adjusted predictor accounts for the attenuation, nor do they establish that health behaviors are unrelated to relational reports.

#### 3.4.2. Sensitivity to MAI straight-lining and influential cases

Because 34 MAI respondents selected the identical response to every item (33 at the scale maximum, one at the minimum), the joint model was re-estimated, and Model B refitted, excluding the 33 ceiling responders, excluding all 34, and excluding the MAI group entirely (Supplementary Table S8). Symptom burden and orienting sensitivity were unaffected or strengthened under every exclusion. The positive distress slope among mothers of infants strengthened when ceiling responders were excluded (β = +0.23, 95% CrI [+0.03, +0.42]) and remained positive but no longer excluded zero when the single floor respondent was also removed (β = +0.15, 95% CrI [−0.05, +0.35]; 93% of posterior mass above zero). Delay discounting was unchanged when ceiling responders were excluded but attenuated to β = −0.088 (p = .036, p-FDR = .105) when the floor respondent was also removed; that respondent, who combined the lowest relational score in the sample with near-maximal discounting, had the largest Cook’s distance of any observation (D = 0.081). The association is therefore small, and its multiplicity-corrected significance in the joint model depends on one case; it is supported, however, by Huber robust regression, which downweights rather than excludes influential observations (β = −0.093, p = .021), by 2,000 bootstrap resamples (95% CI [−0.216, −0.010]), and by its zero-order association, which is unaffected (β = −0.24, p < .001).

### 3.5. Exploratory: Lower-Order Temperament Scales

Orienting sensitivity was the only temperament factor that retained an independent association in the joint model, and its three constituent subscales were examined to locate the effect. This analysis was prompted by the pattern of results rather than planned, and is reported as exploratory.

The association is carried entirely by neutral perceptual sensitivity, the propensity to detect faint external stimulation. Adjusting for symptom burden and group, neutral perceptual sensitivity showed β = +0.203, p < .001, while affective perceptual sensitivity showed β = +0.093 (p = .012) and associative sensitivity showed no association (β = +0.010, p = .805). Entered together, only neutral perceptual sensitivity remained (β = +0.207, p < .001), with associative sensitivity trending negative (β = −0.084, p = .064). Screening all thirteen lower-order ATQ scales on the same basis, neutral perceptual sensitivity was the strongest of the set and survived Bonferroni correction across all thirteen (p < .001; **Table 5**).

**Table 5.** All thirteen lower-order Adult Temperament Questionnaire scales as correlates of relational scores, adjusted for symptom burden and group (exploratory).

| **Subscale** | **β** | **p** | **p-FDR** | **p-Bonferroni** |
|---|---|---|---|---|
| Neutral Perceptual Sensitivity | +0.203 | <.001 | <.001 | <.001 |
| Attentional Control | +0.172 | <.001 | <.001 | <.001 |
| Activation Control | +0.112 | .005 | .022 | .065 |
| Discomfort | −0.104 | .008 | .024 | .103 |
| Fear | −0.099 | .009 | .024 | .122 |
| Affective Perceptual Sensitivity | +0.093 | .012 | .026 | .156 |
| Frustration | −0.089 | .017 | .032 | .224 |
| Inhibitory Control | +0.065 | .093 | .151 | 1.000 |
| High Intensity Pleasure | +0.061 | .105 | .151 | 1.000 |
| Positive Affect | +0.053 | .160 | .209 | 1.000 |
| Sociability | +0.037 | .317 | .375 | 1.000 |
| Sadness | +0.027 | .478 | .518 | 1.000 |
| Associative Sensitivity | +0.010 | .805 | .805 | 1.000 |

*Note.* Each subscale was entered in a separate model adjusting for symptom burden and instrument group. These analyses were not planned and are reported as exploratory.

Its discriminant pattern is notable: neutral perceptual sensitivity correlated .02 with symptom burden, .03 with mindfulness, −.03 with delay discounting, .04 with perceived stress, and .06 with effortful control. It is not redundant with any other construct examined, including mindfulness, despite both concerning attention to present experience. The association was significant only among mothers of children aged 3–12 (β = +0.15, p = .002) and trended among mothers of adolescents (p = .088), and was absent among pregnant women, mothers of infants, and mothers of adult children. Within-factor inconsistency and group specificity both argue for caution, and this finding requires replication before it is built upon.

### 3.6. Contrast: Reports About One’s Own Parents

A separate group of 214 women without children reported on the relationship with their own parents using the Parent–Adult Child Relationship Questionnaire. Role, target relationship, instrument, and context all differ from the maternal-report groups, so this comparison can test whether the observed association differs between the two but cannot isolate which component is responsible.

The estimated symptom–relationship association was weaker in this group, although the comparison is difficult to interpret because report type, target relationship, instrument, and instrument reliability differed simultaneously. In a model containing both report types, the difference in the symptom burden slope (maternal minus own-parent) was −0.266, 95% CI [−0.465, −0.066], p = .009. The corresponding difference for distress was −0.027, [−0.200, 0.146], p = .759, providing no evidence of a difference. Simple slopes were −0.34 (p < .001) for maternal reports and −0.08 (p = .394) for own-parent reports.

The profile association showed the same pattern. Profiles were estimated exclusively within the maternal-report groups. Of the 214 own-parent participants, 213 had complete data on all seven indicators, which were standardized using the means and standard deviations from the maternal group and were assigned based on the trained model, resulting in a mean maximum posterior probability of .936. Assignment was modal and classification uncertainty was not propagated into the interaction test, which should therefore be read as approximate. The profile by report-type interaction was significant, χ²(2) = 14.05, p < .001: profile means ran −0.87, −0.03, and +0.34 in the maternal-report groups and −0.05, −0.07, and +0.14 in the own-parent group.

## 4.0 DISCUSSION

Maternal mental illness is among the most common complications of childbirth, with consequences extending to the child through the quality of the relationship between them(Gavin et al. 2005; Madigan et al. 2018). We approached that relationship as a sustained demand on a mother’s regulatory capacities, and asked, across five family contexts spanning pregnancy through adult children, which of twelve psychological constructs carried independent information about how she described it. The three predictions we derived from that framing were supported. Nearly every construct we examined was associated with relational reports when examined independently, as the existing literature would predict, but when they were combined only three retained independent associations in the pooled model: cross-domain psychiatric symptom burden, delay discounting, and orienting sensitivity. The affective measures on which this literature overwhelmingly relies — depression, anxiety, anger, perceived stress, and negative affect — did not explain additional variance after adjustment for the cross-domain symptom composite and the other measured constructs. A person-centered analysis reached the same conclusion through a different analytic route, as the framing anticipated. Of the three profiles we identified, the one reporting the poorest relational quality was distinguished by symptom burden across domains rather than by elevated stress or negative emotionality. Below we discuss each of these findings in turn, the ways in which the associations varied across family contexts, and what we take to be their implications for how maternal relational functioning is measured and screened, addressing the construct heterogeneity, developmental range, measurement, and analytic limitations identified above.

### 4.1. Relational reports are related to cross-domain symptom burden rather than to specific affective symptoms

Greater breadth of psychiatric symptom burden was related to more negative relational reports in a pooled estimate across five family contexts, from pregnancy through adult children. That pooled estimate did not depend on analytic choices, though the context-specific slopes varied and were credibly negative in the two largest child-age groups rather than in all five. The measure that carried this association was a composite of eight DSM-5 cross-cutting domains(American Psychiatric Association n.d.) — somatic symptoms, suicidal ideation, psychosis, memory, repetitive thoughts and behaviors, dissociation, mania, and substance use — and contains no depression, anxiety, or anger items. Those three domains were entered as separate predictors, were each associated with relational reports on their own, and retained nothing once the composite was included. In other words, what tracked with how a woman described the relationship was not which symptoms she endorsed but how widely they ran, with within-domain severity adding modestly to that. This is the pattern our first two predictions anticipated: the affective measures index a shared liability from overlapping angles, and a measure of that liability’s breadth captures depletion the affective scales do not.

In line with our findings, a general-factor account of psychopathology would predict exactly this pattern. Across twenty years of the Dunedin cohort, psychiatric disorder was explained better by a single general dimension than by separate internalizing, externalizing, and thought-disorder factors, and higher standing on that dimension was associated with greater life impairment, greater familiality, worse developmental histories, and more compromised early-life brain function(Caspi et al. 2014). A decade of subsequent work indicates that the reliable variance in psychopathology measures sits largely in this general factor, with the residual specific factors poorly measured(Caspi et al. 2024). This non-specificity is now understood to account for why distinct diagnoses share risk factors, biomarkers, and treatment response(Caspi and Moffitt 2018). We hypothesize, without claiming equivalence between a cross-sectional composite and a longitudinally estimated latent factor, that a liability of this general kind is what operates on the mother–child relationship, and that the well-replicated finding that maternal depression, maternal anxiety, and maternal stress are each related to poorer bonding(O’Dea et al. 2023) reflects three views of that shared dimension rather than three separable contributions. Mechanistically, this would make sense. The dyadic processes through which maternal state reaches the child — coordination of gaze, vocalization, and affect, and the co-regulation of heart rate, cortisol, and autonomic arousal(Bell 2020; Feldman 2007) — are demanding of attention, energy, and emotional availability, and we would not expect them to be disrupted only by symptoms that fall within the mood domain. A mother who is not sleeping, who is experiencing intrusive thoughts, or who is using substances has fewer resources available for those exchanges regardless of whether she meets criteria for a depressive or anxious disorder(A. Stein et al. 2014).

Two interpretive constraints apply. First, substantial shared variance among the affective measures is not the same as identity, and the absence of a detectable unique contribution is not evidence that no unique contribution exists; with predictors this highly correlated, our power to resolve small independent effects is limited. Second, these data are cross-sectional. We can say that symptom breadth and relational report covary, but not that symptoms degrade the relationship, that a difficult relationship elevates symptoms, or that some third factor produces both. Prospective work with repeated assessment of both would be needed to separate these possibilities.

### 4.2. Relational reports are related to delay discounting, a behavioral measure of future valuation

Our third prediction was that constructs indexing regulatory capacity, rather than its depletion, would retain associations independent of symptom measures. Of the twelve constructs we examined, only one was a behavioral task rather than a self-report questionnaire, and it was among the three that survived adjustment. Women who discounted delayed rewards more steeply described their relationships more negatively. To our knowledge, this is the first peer-reviewed study to connect delay discounting to maternal reports of relationship quality. Delay discounting indexes the balance between the impulsive and executive systems that govern intertemporal choice(Bickel et al. 2018), and it has been identified as a trans-disease process spanning substance use disorder, major depressive disorder, schizophrenia, borderline personality disorder, bipolar disorder, and a range of maladaptive health behaviors(Amlung et al. 2019; Bickel et al. 2019). We have previously shown that it is also linked to dissociation and other trauma symptomatology(Basso et al. 2023). The present finding extends that body of work into the relational domain, and does so in a population in which decision-making has rarely been measured at all.

Interestingly, delay discounting behaved here in precisely the manner its trans-disease characterization would predict. It was close to unrelated to the affective measures, and its direction was the same in four of the five family contexts, a pattern consistent with, but not establishing, a behavioral process partly distinct from current affective state. We speculate that steep discounting bears on the mother–child relationship through the temporal structure of caregiving itself. Caregiving often asks a mother to repeatedly choose a delayed, diffuse reward over an immediate one: staying with a crying infant, holding a boundary with a toddler, tolerating an adolescent’s hostility without withdrawing. A narrowed temporal window would make these choices more costly(Bickel et al. 2018), and over thousands of repetitions we would expect that cost to register in how a woman describes the relationship. This account is speculative and the present design cannot test it, but it is directly testable in a design that pairs a discounting task with observed caregiving behavior.

The association was negative in four of the five family contexts and of similar magnitude in three of them, and it varied less across contexts than any other predictor we examined, with only the adolescent group running slightly positive. It reached credibility in two groups, with the remaining intervals too wide to resolve, which is unsurprising given that three of our five groups contained fewer than 100 women. We therefore regard the evidence for discounting as consistent in direction but less precisely estimated than that for symptom burden, and we note that the pooled estimate and the group-specific estimates answer different questions. It is also sensitive to one influential respondent in the infant group, though robust estimation and resampling, which address influence without excluding cases, support it. The point estimate was largest among pregnant women, but that group was small and its slope was not shown to differ from the others, so we draw no conclusion specific to pregnancy and note only that a prenatal test is worth conducting directly. The clinical interest of this finding lies in the fact that discounting is modifiable. Episodic future thinking and related interventions reduce discounting rates in several populations(J. S. Stein et al. 2018), and the task used here takes only a few minutes to administer(Koffarnus and Bickel 2014).

### 4.3. Relational reports are related to neutral perceptual sensitivity, a temperamental measure of stimulus detection

Orienting sensitivity was the second capacity measure named in our third prediction. Of the four higher-order temperament factors measured by the Adult Temperament Questionnaire(Evans and Rothbart 2007), it alone retained an independent association, and decomposing it showed that the association was carried entirely by one of its three subscales. Neutral perceptual sensitivity, the propensity to detect slight, low-intensity stimulation in the environment, was the strongest of all thirteen lower-order scales and survived Bonferroni correction across them. The association was positive: women higher on this trait described their relationship with their child more favorably, though the opposite direction would also have been defensible a priori, as a heightened propensity to register faint stimulation might equally have been expected to make the sensory demands of caregiving more taxing. The finding was also independent of everything else we measured. Where nearly every other construct collapsed into cross-domain symptom burden once entered jointly, neutral perceptual sensitivity was essentially uncorrelated with it, with perceived stress, and with delay discounting. It was likewise uncorrelated with mindfulness(Brown and Ryan 2003), despite both constructs concerning attention to present experience. Effortful control, the other temperamental capacity we had expected to retain an association, did not; it was substantially correlated with the affective measures and its variance was absorbed by them, which is itself consistent with the first prediction.

We hypothesize that this points toward a temperamental substrate for maternal sensitivity, the capacity to notice and respond appropriately to a child’s subtle cues, which is the central explanatory mechanism in attachment theory and the most reliable target of intervention(Bakermans-Kranenburg et al. 2003). Sensitivity is ordinarily established by observing a mother with her child; the measure used here asked only about the detection of faint stimulation in general, with no reference to children or to caregiving, and it nonetheless related to how a woman described the relationship. We therefore speculate that the capacity to register low-intensity cues may be partly temperamental, and that women differ in it before and independently of anything specific to motherhood. This analysis was exploratory, driven by the pattern of results rather than predicted in advance, and the association varied across contexts. It was positive among mothers of children aged 3 to 12 and of adolescents, with similar slope magnitudes in both, reaching credibility in the former and trending in the latter. It was absent among mothers of infants, pregnant women, and mothers of adult children, the latter two groups being too small to resolve an association of this magnitude in either direction. We speculate that this localization is itself informative, as mid-childhood and adolescence may be the periods in which a mother’s read on her child depends most on detecting subtle cues rather than on overt signals.

### 4.4. Psychological profiles differ in relational reports

Nearly all of the work in this area is variable-centered, estimating the average association between one predictor and a relational outcome, yet psychological characteristics co-occur within individuals. Person-centered methods have been applied in the perinatal period to bonding trajectories(de Waal et al. 2026) and to profiles of co-occurring maternal mental-health symptoms(Kingston et al. 2018), but not to the broader set of psychological characteristics that may accompany relational difficulty. We identified three profiles, and as hypothesized, they differed in relational reports in the expected ordering. Importantly, however, the women who reported the greatest difficulty were not those characterized by elevated stress and negative emotionality, which formed a profile of their own and which are the characteristics this literature most often measures. They were the women whose psychiatric symptoms ran across multiple domains and were accompanied by elevated medical burden, poor sleep, and low effortful control. The ordering across profiles was large, and it held after accounting for family context, under the Bolck–Croon–Hagenaars correction for classification error, and in a held-out subsample, although at roughly half the magnitude there.

That the variable-centered and person-centered analyses converge on the same characterization strengthens both, as the two approaches are vulnerable to different artifacts: a regression coefficient can be driven by a minority of extreme cases, while a profile solution can impose an arbitrary cut through an underlying continuum. We also note that profile membership varied markedly by family context, with the symptomatic profile describing 51.3% of pregnant women and none of the mothers of adult children. This is consistent with what is known about the concentration of psychiatric morbidity during the perinatal period(Gavin et al. 2005), though our pregnancy group was small and these proportions should not be read as prevalence estimates. Because the profiles were both derived and evaluated within this dataset, the hold-out analysis constitutes an internal robustness check rather than independent confirmation, and replication in a separate sample is needed.

### 4.5. The symptom–relationship association varies across family contexts

Most of what is known about maternal psychological functioning and the mother–child relationship comes from pregnancy and the first postpartum year, even though fewer than half of maternal depressive episodes begin postpartum(Wisner et al. 2013) and depressive symptoms remain as prevalent at four years postpartum as in the first year(Woolhouse et al. 2015), and the work that does extend further is sparse and uses heterogeneous outcome definitions(Badovinac et al. 2018; Hale et al. 2020; Risi et al. 2021). Our design was intended to span that range, and the associations we observed were not uniform across it. In one context they inverted: among mothers of children under two, higher distress was associated with higher reported attachment, while every older group showed the opposite. This reversal persisted under partial pooling, a procedure that works against extreme group estimates.

We cannot determine whether this reflects something about early motherhood or something about the instrument administered in that group. The Maternal Attachment Inventory contains no items concerning conflict or difficulty, so a mother who is struggling has no means of reporting that struggle on it, and it produced a response pattern that the other instruments did not. The ceiling effect itself does not account for the reversal: excluding the respondents at the scale maximum strengthened rather than weakened the positive distress slope (Supplementary Table S8), although the interval widened to include zero once the single floor respondent was also excluded. What remains is the instrument’s item content, already noted, and the possibility that the early postpartum period is one in which distress and intense attachment genuinely co-occur, as a mother who is anxious about her infant may also be highly preoccupied with that infant. The comparison between the adolescent RIPA and its adult-child adaptation holds the instrument family more nearly constant, but not the instrument itself, and showed no difference, which is consistent with a measurement explanation but is not sufficient to establish it. Until the reversal is replicated with a common instrument administered across contexts, we regard it as an observation rather than as evidence about the psychology of early motherhood.

This is the central interpretive limitation of our design. Because each family context was assessed with a different instrument, context and measurement cannot be separated. Reviews of parent-report relational measures find uneven psychometric support across the available instruments(Wittkowski et al. 2020), and the three constructs that an umbrella term such as attachment is made to cover — a mother’s feelings toward her fetus or infant, a child’s behavior toward a caregiver, and a parent’s account of closeness with an older child — are related but not equivalent(Bicking Kinsey and Hupcey 2013). For the same reason, the robustness of the pooled association should be read as evidence that it does not depend on analytic choices, rather than as evidence that it holds within every family context.

### 4.6. The symptom–relationship association is weaker in reports about one’s own parents

Women reporting on their own parents rather than on a child showed a markedly weaker relation between symptom burden and relationship quality, and no profile difference at all. The attenuation was specific to symptom burden; distress behaved similarly in both groups. We offer two interpretations. One possibility is that the difference lies in the reporting rather than in the relationships. A woman’s current state may color how she describes a relationship she is actively conducting, with its daily demands and frictions, while her account of a relationship formed decades earlier has settled into a more stable narrative that current mood does not disturb. The other possibility is that the difference is real: current psychological functioning can act on a relationship that is still being built, but not on one already formed.

Our design cannot separate these, as role, target, instrument, and life stage all differ simultaneously between the comparison groups. We note that measurement unreliability in either relational instrument would attenuate the observed association, and the difference between groups may be partly attributable to differences in instrument reliability rather than to reporting dynamics. If the second reading is correct, it would suggest that current psychological functioning acts on relationships that are actively being conducted, which is precisely the circumstance in which the associations reported here were observed. What the contrast does establish is that the same symptom measure did not relate to relational report in the same way across reporting situations, which is in line with evidence that informant and method shape which associations emerge: in a cohort followed from pregnancy to three months postpartum, prenatal anxiety disorders predicted self-reported bonding impairment but not observed interaction quality, while depressive symptoms predicted lower observed sensitivity(Nath et al. 2019), suggesting that self-reported and observed measures may capture different aspects of maternal functioning rather than one being a degraded version of the other. We take this as a caution against treating self-reported relational scores as interchangeable across reporting contexts, and as further reason to pair self-report with observational measurement in work of this kind.

### 4.7. Implications for measurement and screening

The clearest implication of our findings concerns what is worth measuring. Eleven of the twelve constructs we examined were associated with relational reports individually, yet only three retained a detectable unique association once entered together. Adding a further self-report mood scale to models already containing depression, anxiety, and stress is therefore unlikely to generate a new finding; it is likely to generate the same finding again. The measures that did add something were of a different kind — a behavioral task and a temperamental dimension outside the negative-affect domain — which is what a resource account predicts: measures of the capacities caregiving draws on carry information that measures of their depletion do not. We would therefore encourage future work in this area to invest in behavioral, temperamental, and observational measurement rather than in additional affective questionnaires. We emphasize that this is not an argument that depression and anxiety are unimportant to the mother–child relationship; the mechanistic literature on dyadic co-regulation makes that implausible(Feldman 2007). It is an argument that measuring them in several ways does not yield unique findings.

A second implication concerns the relational outcome itself. The instruments available across the developmental range are not interchangeable, the adaptation we used for adult children performed poorly, and one instrument offered mothers no way to report difficulty. Extending this literature beyond infancy will require either a relational measure validated across developmental stages or an explicit strategy for modeling non-equivalence, and we would regard the development of such a measure as a priority for the field.

Finally, we raise a possibility for screening, which we offer as a hypothesis for prospective testing rather than as a recommendation. Detection and treatment of maternal mental illness remain poor at every step from screening through diagnosis to remission(Cox et al. 2016), mental health conditions have overtaken hemorrhage and cardiac conditions as the leading underlying cause of pregnancy-related death(Trost et al. 2022), and the proportion of US mothers rating their mental health as fair or poor rose by nearly 65% between 2016 and 2023(Daw et al. 2025). Screening in this setting is organized almost entirely around perinatal mood and the first postpartum year, yet only 40.1% of depressive episodes identified postpartum had begun in that period(Wisner et al. 2013), depressive symptoms are as prevalent at four years postpartum as at any point in the first year(Woolhouse et al. 2015), and the dimension that tracked relational report in our data ran across domains that mood-focused screening does not assess. Given that offspring of mothers with perinatal depression show elevated odds of depression, disruptive behavioral symptoms, and socioemotional difficulties into adulthood(Madigan et al. 2018; Tirumalaraju et al. 2020; Tusa et al. 2025), we consider it worth determining whether a brief cross-cutting symptom measure identifies mothers at risk of relational difficulty whom mood screening currently misses. The DSM-5 Level 1 measure was developed to inform dimensional assessment rather than as a screening instrument, and its domain thresholds show strong sensitivity but limited specificity in community samples(Doss and Lowmaster 2022), so any such application would require validation against outcomes in a perinatal population.

### 4.8. Limitations and future directions

This study has several limitations that warrant consideration. All measures were collected at a single time point, so no temporal or causal claims are supported. Relational outcomes and most predictors were self-reported, which leaves the associations among them open to shared-method variance; delay discounting was our only task-based measure. The instrument was perfectly confounded with family context, so differences between contexts cannot be attributed to the child’s age. The relational instruments measure related but non-equivalent constructs, and the adult-child adaptation showed poor internal consistency (α = .44) and has no published validation; the comparison between maternal and own-parent reports in Section 3.6 should accordingly be read as descriptive. Our pregnancy group and our group of mothers of adult children were small. Among highly correlated predictors, a failure to detect a unique contribution should not be read as evidence that none exists. The latent profiles were derived and evaluated within the same dataset. The profile indicator set was fixed before the temperament decomposition in Section 3.5 and therefore does not include orienting sensitivity; whether a profile defined partly by that trait would emerge is a question for replication. The medical history checklist was unreliable at the upper end of its range (Section 3.3.3). Five analytic decisions were made after inspecting the data and are enumerated in Section 2.4.1; our specification curve quantifies analytic flexibility for the focal association but not for the exploratory analyses, including the temperament decomposition reported in Section 4.3.

Additionally, our sample was recruited online through Amazon Mechanical Turk. We applied several procedures to address the data quality concerns associated with crowdsourced samples, excluding spam-flagged responses, incomplete submissions, respondents failing attention checks, and duplicate submissions by the same worker, which together reduced 1,502 raw responses to the 840 retained here. We note that these data were collected in 2021, prior to the period in which substantial deterioration in crowdsourced data quality has been documented(Webb and Tangney 2024), and that the internal consistencies observed here (α = .86 to .95) and the 60-day test–retest correlation for the discounting task (r = .64) are not consistent with widespread inattentive or automated responding. Nonetheless, the sample was predominantly White, educated, and partnered, which limits generalization and may restrict the observed range of both symptom burden and relational difficulty.

Several directions follow from this work. Most pressingly, observational measurement of caregiving behavior should be paired with the self-report instruments used here, both to test whether the symptom–relationship association holds for observed interaction and to test our specific hypothesis that neutral perceptual sensitivity predicts observed maternal sensitivity. The associations reported here should also be examined prospectively, with repeated assessment of symptoms and relational quality, in order to establish direction of effect. A relational instrument administered unchanged across the developmental range would allow the group differences we observed, and particularly the reversal among mothers of infants, to be separated from measurement artifact. We would also encourage interventional work: because delay discounting can be reduced through episodic future thinking(J. S. Stein et al. 2018), a trial testing whether such training alters maternal relational outcomes would convert a correlational finding into a causal test. Finally, the dyadic processes through which maternal state is thought to reach the child are measurable at the level of autonomic and neuroendocrine synchrony between mother and infant(Bell 2020), and hyperscanning approaches now permit the simultaneous measurement of neural activity in both members of a dyad during interaction(Culligan et al. 2025). Because the constructs we examined here were measured in the mother alone, while the outcome of interest is a relationship, applying such methods to mother–child dyads would allow the associations reported here to be tested at the level at which they are presumed to operate.

### 4.9. Conclusion

We began from the observation that maternal mental illness is common, under-detected, and consequential for children through the relationship between mother and child, and that the evidence on that relationship rests on a narrow set of overlapping mood measures collected almost entirely in the perinatal year. Treating caregiving as a sustained demand on a mother’s regulatory capacities, and psychiatric liability as what depletes them, we predicted that the mood measures would prove largely redundant with one another, that breadth of symptoms would carry information the mood measures do not, and that measures of the capacities themselves would carry information independent of symptoms. Each prediction was supported in a pooled estimate across family contexts spanning pregnancy through adult children. How a woman described her relationship with her fetus or child was related not to which psychiatric symptoms she reported but to how widely they ran; the affective measures on which this literature relies showed no detectable unique association once that breadth was accounted for; a person-centered analysis arrived at the same characterization independently; and delay discounting, a brief behavioral measure of future valuation, was related to relational reports for the first time in a peer-reviewed study, with an exploratory temperamental correlate reported as a hypothesis. These are cross-sectional associations across non-equivalent relational instruments, and they identify candidate correlates rather than mechanisms. What they suggest is that the psychological dimension most closely tied to maternal relational reports is broader than the perinatal mood constructs around which both research and screening are currently organized, and that the next gains in understanding this relationship are more likely to come from measuring the capacities caregiving draws on than from measuring mood one more way. Because one of those capacities, future valuation, is modifiable through episodic future thinking, that proposition can be tested directly.

## Data Availability

All data produced in the present study are available upon reasonable request to the authors

