## Supplementary Information for "Beyond Perinatal Mood: Breadth of Psychiatric Symptoms and Delay Discounting as Correlates of Mothers’ Relational Reports from Pregnancy Through Adult Children"

**Supplementary Methods**

**S1. Relational instruments**

**Prenatal Attachment Inventory (PAI).**

The PAI measures the affectionate relationship that develops between a woman and her fetus. It comprises 21 items rated on a four-point scale, summed to a total ranging from 21 to 84, with higher scores indicating greater prenatal attachment. Published alphas range from .85 to .89; in the present sample, α = .91 (n = 39). This value is at the upper end of the published range.

**Maternal Attachment Inventory (MAI).**

The MAI measures maternal affectionate attachment and is used to assess a mother’s emotional response to her infant (validated for children 0 to 2 years). Items are rated on a four-point scale (almost never to almost always) and summed. The published alpha is .85; in the present sample, α = .98 (n = 145). This unusually high value reflects a pronounced ceiling effect: 34 of 145 mothers (23.4%) selected the identical response option for every item, 33 of them the maximum. All MAI items are positively worded and the instrument contains no items assessing conflict or difficulty, which plausibly accounts for both the ceiling effect and the inflated reliability estimate.

**Child–Parent Relationship Scale (CPRS).**

The CPRS measures parent perceptions of the relationship with their child (validated for children 3–12 years). Items are rated on a five-point scale and summed into three subscales: conflict, positive aspects of the relationship, and dependence. Published subscale alphas are .83, .72, and .50 respectively. In the present sample, α = .80 across all items (n = 298); because the subscales are scored in opposing directions, the item-level total is reported for reference only.

**Revised Inventory of Parental Attachment (RIPA).**

Derived from the Inventory of Parent and Peer Attachment, the RIPA was adapted so that parents report on their attachment relationship with their child (validated for children 13–17 years). It comprises 30 items rated on a five-point scale. The published validation (N = 212 parents, predominantly mothers) reported α = .91 for trust/avoidance and α = .72 for communication. The version administered to parents of children aged 18 and over is an adaptation for which no separate published validation exists. In the present sample, α = .75 for the adolescent version (n = 91) and α = .44 for the adult-child version (n = 40); the low reliability of the adult-child version, together with its unvalidated status and small subsample, warrants considerable caution.

**Parent–Adult Child Relationship Questionnaire (PACRQ).**

Administered to the 214 women without children, who reported on the relationship with their own parents, and used only in the contrast analysis of Section 3.6. Factors of regard, responsibility, and control were derived separately for mothers and fathers. Published alphas are .87 (mother regard), .82 (mother responsibility), .86 (father regard), .74 (father responsibility), and .87 (father control). In the present sample, α = .86 for the mother items (n = 211) and α = .70 for the father items (n = 171). The score used Mother Regard and Father Regard only; Responsibility and Control index filial obligation and parental dominance rather than low attachment, and the reverse-scored Mother Responsibility subscale correlated negatively with Mother Regard (r = −.22). A sensitivity variable retaining all five subscales was also computed and gave the same pattern of results.

**S2. Psychological and health measures**

**DSM-5 Level 1 Cross-Cutting Symptom Measure.**

Twenty-three items rated 0 (not at all), 1 (rare, less than a day or two), 2 (several days), 3 (more than half the days), and 4 (nearly every day) cover 13 domains. The 13 domains encompass various psychiatric dimensions as follows: depression, anger, mania, anxiety, somatic symptoms, suicidal ideation, psychosis, sleep problems, memory, repetitive thoughts and behaviors, dissociation, personality functioning, and substance use. Domains were scored using the standard highest-item-within-domain rule. The measure is designed for domain-level rather than total scoring, and the DSM-5 field trials therefore report domain-level internal consistency; validation studies of translated adult versions report total-score alphas of .94 and .95. In the present sample, α = .95, consistent with published values.

**Other psychological measures.**

Mindful Attention Awareness Scale (15 items, α = .93); Perceived Stress Scale (10 items, α = .89); Adult Temperament Questionnaire short form (77 items, α = .91), yielding four higher-order factors and thirteen lower-order scales; Autism Spectrum Quotient (50 items, α = .86), summed and used dimensionally, with the conventional binary threshold of 26 not applicable to this scoring method.

**Health measures.**

Pittsburgh Sleep Quality Index; a 41-item medical history checklist (American Health Information Management Association, My Personal Health Record, Adult PHR Form 2006, Section F) scored as a count of endorsed conditions; the Stanford Brief Activity Survey, classifying activity from 1 to 5 via a lookup matrix crossing on-the-job and leisure-time activity; a sitting/sedentary behavior protocol assessing four domains separately for weekdays and weekend days, top-coded at "8 or more" hours, which 254 participants (30.7%) selected for at least one domain; and the Block rapid food screeners for dietary fat (17 items) and fruit and vegetable intake (7 items). Body mass index was computed from self-reported height and weight.

**S3. Test–retest reliability**

Fifty-eight workers completed the survey in both collection waves, providing an incidental test–retest sample with a median interval of 60 days (range 59–68). Stability was good for most measures (median r = .78), and no measure showed a significant mean change between administrations (all p > .11), indicating no systematic practice or regression effect.

**Supplementary Table S1.** *Test–retest reliability over a median interval of 60 days (n = 58).*

| **Measure** | **r** | **ρ** | **ICC(2,1)** |
| --- | --- | --- | --- |
| DSM-5 Cross-Cutting total | .92 | .90 | .92 |
| Autism Spectrum Quotient | .91 | .84 | .91 |
| Perceived Stress Scale | .90 | .86 | .89 |
| ATQ Effortful Control | .88 | .88 | .88 |
| ATQ Extraversion/Surgency | .87 | .84 | .86 |
| ATQ Negative Affect | .81 | .75 | .81 |
| ATQ Orienting Sensitivity | .74 | .75 | .73 |
| Fruit and vegetable intake | .72 | .74 | .69 |
| Dietary fat intake | .71 | .76 | .71 |
| Mindful Attention Awareness Scale | .65 | .70 | .65 |
| Delay discounting | .64 | .67 | .64 |
| Medical history (condition count) | .03 | .60 | .02 |

*Note.* N = 58 for every measure. The medical history checklist is the exception to an otherwise acceptable pattern: three participants endorsed 39, 39, and 18 of 41 conditions at one administration and none at the other, which eliminates the product–moment correlation while leaving rank-order agreement intact. This measure should be regarded as unreliable at the upper end of its range.

**Supplementary Results**

**S4. Group-specific credible intervals, Model A**

**Supplementary Table S2.** *Group-specific standardized slopes with 95% credible intervals from Model A (N = 613).*

| **Group** | **n** | **Symptom burden** | **Delay discounting** | **Orienting sensitivity** |
| --- | --- | --- | --- | --- |
| Pregnant | 39 | +0.09 [−0.15, +0.31] | −0.19 [−0.39, −0.02] | −0.05 [−0.43, +0.18] |
| Child 0–2 | 145 | −0.44 [−0.58, −0.31] | −0.11 [−0.23, +0.01] | +0.02 [−0.13, +0.15] |
| Child 3–12 | 298 | −0.48 [−0.60, −0.36] | −0.11 [−0.22, −0.00] | +0.10 [+0.01, +0.19] |
| Child 13–17 | 91 | −0.24 [−0.47, +0.01] | +0.07 [−0.14, +0.33] | +0.11 [−0.03, +0.27] |
| Child 18+ | 40 | −0.32 [−0.68, +0.05] | −0.12 [−0.44, +0.14] | −0.03 [−0.31, +0.15] |
| Population mean |  | −0.28 [−0.39, −0.17] | −0.09 [−0.19, +0.01] | +0.04 [−0.09, +0.13] |
| SD across groups |  | 0.28 | 0.16 | 0.13 |

*Note.* Posterior means with 95% credible intervals. Fitted with target acceptance of .99: maximum R-hat 1.002, minimum bulk effective sample size 3,252, zero divergent transitions. With only five group levels, population-level summaries should be read as describing these observed, measure-defined contexts rather than as estimates for a broader population of contexts.

**S5. Latent class enumeration**

**Supplementary Table S3.** *Latent profile class enumeration (N = 611).*

| **Classes** | **Log-likelihood** | **BIC** | **LRT vs. k−1** | **BLRT p** | **Entropy** | **Smallest class** |
| --- | --- | --- | --- | --- | --- | --- |
| 2 | −5132.7 | 10451.5 | 1865.2 | .010 | .866 | 44.8% |
| 3 | −4844.1 | 9970.4 | 577.3 | .010 | .860 | 15.7% |
| 4 | −4684.9 | 9748.2 | 318.4 | .010 | .831 | 12.3% |
| 5 | −3881.9 | 8238.5 | 1605.9 | .010 | .911 | 5.6% |
| 6 | −3824.9 | 8220.7 | 114.1 | .010 | .894 | 5.2% |

*Note.* BIC declined monotonically and the bootstrapped likelihood ratio test remained significant through six classes, a known behavior of both indices at large sample sizes. Class selection therefore weighted entropy, class size, and interpretability. Entropy was comparable for the two- and three-class solutions (.866, .860) and fell at four (.831); the five- and six-class solutions achieved higher entropy only by splitting off classes containing 5–6% of the sample. The three-class solution was preferred over the two-class solution because the latter absorbed the distressed–symptomatic profile into a broad distressed class (n = 274) with a mean symptom burden of 0.81 SD rather than 1.75, dissolving the high-symptom group that distinguishes breadth of psychiatric symptoms from elevated distress.

**S6. Profile indicator means**

**Supplementary Table S4.** *Standardized indicator means by latent profile (N = 611).*

| **Profile** | **n** | **%** | **Stress** | **Mindfulness** | **Neg. emotion** | **Effortful control** | **Symptoms** | **Sleep** | **Medical burden** |
| --- | --- | --- | --- | --- | --- | --- | --- | --- | --- |
| 1. Distressed–symptomatic | 96 | 15.7 | 0.49 | −0.39 | 0.30 | −0.71 | 1.77 | 0.57 | 1.32 |
| 2. Mildly strained | 282 | 46.2 | 0.50 | −0.34 | 0.43 | −0.35 | 0.06 | 0.42 | −0.12 |
| 3. Resilient | 233 | 38.1 | −0.81 | 0.58 | −0.65 | 0.72 | −0.80 | −0.74 | −0.41 |

*Note.* Values are standardized indicator means. The distressed–symptomatic profile is distinguished primarily by psychiatric symptom burden and medical burden; profiles 1 and 2 show comparable stress and negative emotionality and differ chiefly in symptom burden, medical burden, and effortful control.

**S7. Correction for classification error**

**Supplementary Table S5.** *Profile means before and after correction for classification error (N = 611).*

| **Profile** | **Modal n** | **Naive M** | **BCH M** | **SE** |
| --- | --- | --- | --- | --- |
| Distressed–symptomatic | 96 | −0.86 | −0.88 | 0.11 |
| Mildly strained | 282 | −0.02 | −0.04 | 0.06 |
| Resilient | 233 | +0.38 | +0.40 | 0.05 |

*Note.* Bolck–Croon–Hagenaars three-step correction, weighting cases by the inverse of the empirical classification error matrix. Diagonal entries of the error matrix were .919, .932, and .949. Wald χ²(2) = 150.31, p < .001.

**S8. Health behaviors**

**Supplementary Table S6.** *Health behaviors before and after adjustment for psychological constructs (N = 611).*

| **Health behavior** | **Zero-order r** | **Adjusted β** | **p** |
| --- | --- | --- | --- |
| Medical burden | −.28 | −0.103 | .015 |
| Dietary fat intake | −.23 | −0.068 | .116 |
| Sleep problems (PSQI) | −.19 | +0.059 | .171 |
| Fruit and vegetable intake | −.15 | −0.042 | .312 |
| Sedentary hours per day | −.11 | +0.014 | .721 |
| Physical activity | −.01 | −0.045 | .219 |
| Body mass index | −.00 | −0.008 | .835 |

*Note.* Adjusted estimates are from models containing symptom burden, the distress composite, delay discounting, instrument group, and the health behavior in question. Entered as a block, the seven health behaviors did not improve fit, F(7, 596) = 1.76, p = .094, ΔR² = .016. The sleep coefficient reverses sign on adjustment.

**S9. Confirmatory factor analysis**

A three-factor confirmatory factor analysis was fitted to establish that the predictor set has coherent latent structure. Fit was marginal: CFI = .904, TLI = .884, RMSEA = .089, SRMR = .068. The model is reported as a measurement model only; the composites entered in the analyses are observed means, not factor scores.

**Supplementary Table S7.** *Indicators and standardized loadings for the three-factor confirmatory model.*

| **Factor** | **Indicators** | **Standardized loadings** |
| --- | --- | --- |
| Psychiatric symptoms | DSM-5 domains: dissociation, repetitive thoughts, suicidal ideation, psychosis, memory, mania, somatic symptoms | .65 to .86 |
| Self-regulation | ATQ Attentional Control, Activation Control, Inhibitory Control, MAAS | .56 to .87 |
| Negative emotionality | ATQ Fear, Discomfort, Frustration, Sadness | .52 to .86 |

*Note.* Factor intercorrelations: self-regulation with negative emotionality r = −.61; self-regulation with psychiatric symptoms r = −.55; negative emotionality with psychiatric symptoms r = .33. Mindfulness loads as an indicator of self-regulation rather than constituting a dimension of its own, so effects of the distress composite cannot be attributed to mindfulness specifically.

**S10. Sensitivity analyses**

**Alternative inclusion rules.**

Nine repeat participants completed a different relational instrument at the two administrations, some plausibly reflecting development (an infant passing age two, a new pregnancy) and some not. Refitting the primary model on an alternative sample excluding these cases left all estimates essentially unchanged.

**First versus later submission.**

For the 58 workers who completed the survey twice, substituting the earlier response for the later one left all Model B estimates essentially unchanged (infant-group distress slope +0.21 versus +0.20; infant-group symptom burden slope −0.51 versus −0.51; posterior probability of a positive infant-group distress slope .992 versus .984). Only 35 of the 58 repeat submitters fall within the maternal-report groups, so the substitution affects 35 of 613 cases.

**Race and ethnicity.**

Adding Hispanic ethnicity and White race as population-level covariates left every group-level slope essentially unchanged, and neither covariate was credible. Given the sample’s homogeneity, both are reported in the sample description rather than modeled.

**PACRQ scoring.**

Under the all-five PACRQ scoring, the pattern of results in the own-parent contrast group was unchanged.

**MAI straight-lining and influential cases.**

Thirty-four MAI respondents selected the identical response to every item, 33 at the scale maximum and one at the minimum. The joint model (Table 3) and Model B were re-estimated excluding the 33 ceiling responders, excluding all 34, and excluding the MAI group entirely, with the MAI outcome re-standardized within retained respondents after each exclusion. Huber robust regression and a 2,000-resample bootstrap were applied to the full sample.

**Supplementary Table S8.** *Sensitivity of the joint model and the infant-group distress slope to MAI straight-lining and influential cases.*

| **Analysis** | **N** | **Symptom burden β (p)** | **Delay discounting β (p; p-FDR)** | **Orienting sensitivity β (p; p-FDR)** | **Infant-group distress slope [95% CrI]** |
| --- | --- | --- | --- | --- | --- |
| Full maternal sample | 613 | −0.382 (<.001) | −0.114 (.005; .031) | +0.105 (.009; .038) | +0.21 [+0.04, +0.38] |
| Excluding 33 ceiling responders | 580 | −0.358 (<.001) | −0.117 (.006) | +0.103 (.015) | +0.23 [+0.03, +0.42] |
| Excluding all 34 straight-liners | 579 | −0.381 (<.001) | −0.088 (.036; .105) | +0.128 (.002; .014) | +0.15 [−0.05, +0.35] |
| Excluding MAI group | 468 | −0.210 (.003) | −0.077 (.094) | +0.124 (.010) | — |
| Huber robust regression, full sample | 613 | −0.390 | −0.093 (.021) | +0.129 (.001) | — |
| Bootstrap 95% CI, full sample | 613 | — | [−0.216, −0.010] | — | — |

*Note.* Joint-model coefficients are standardized. p-FDR is Benjamini–Hochberg across the twelve joint-model tests, shown where the exclusion changed the correction outcome. Infant-group distress slopes are from Model B. The floor respondent combined the lowest relational score in the sample with near-maximal delay discounting and had the largest Cook’s distance of any observation in the joint model (D = 0.081); 98.6% of bootstrap resamples returned a negative delay discounting coefficient. Joint-model rows use the 613 maternal respondents with a relational score. Model B rows use the same 613 respondents.

**Supplementary Figures**

**Supplementary Figure S1.** *Specification curve with analytic choices, for the 144 specifications summarized in Figure 4.*

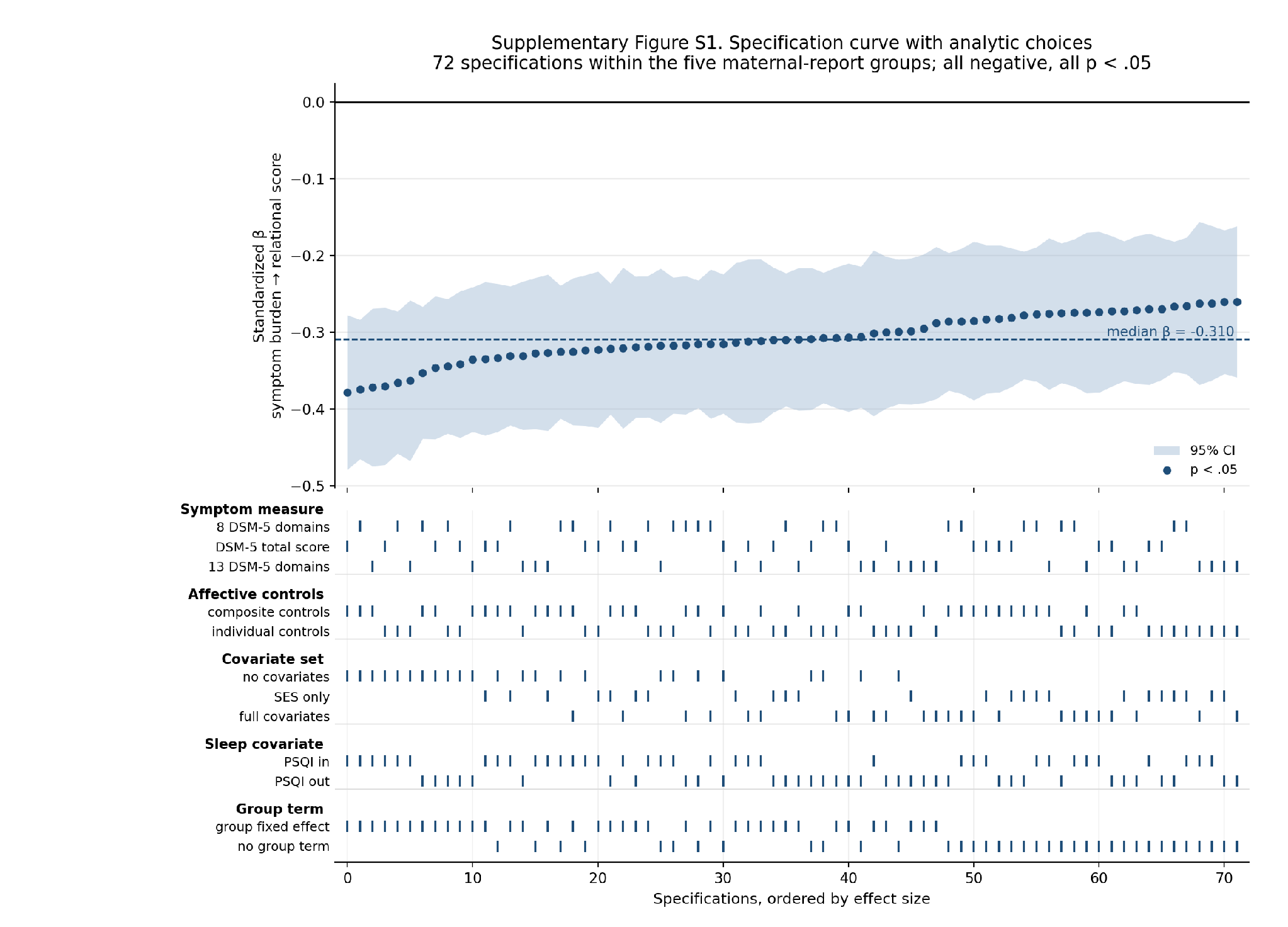

*Note.* Upper panel: standardized estimates from 144 analytic specifications within the five maternal-report groups, ordered by effect size, with 95% confidence intervals. All 144 were negative and all reached p < .05. The dashed line marks the median estimate. Lower panel: the analytic choice producing each estimate, with ticks indicating which option was used. Group inclusion was not varied, because adding the own-parent group changes the relational construct under study rather than the analytic specification of a single estimand.
